# Dysregulations in Cardiogenic Mechanisms by TGF-beta and Angiotensin II in Cardiac Remodeling Post-Ischemic Injury: a systematic review

**DOI:** 10.1101/2024.07.11.24310260

**Authors:** Ovais Shafi, Kashaf Zahra, Haider Hussain Shah

## Abstract

**Objective:** The objective of this study is to look into how TGF-beta and Angiotensin II disrupt cardiogenic regulators (Isl1, Brg1/Baf60-Smarcd3 complex, Nkx2-5, GATA4, Tbx5, Mef2c, HAND1/2, MYOCD, MSX2, HOPX, Wnt-signaling pathway, Notch, FGF, BMPs) during cardiac remodeling post-ischemic injury.

**Background:** Cardiac remodeling post-ischemic injury, influenced by TGF-beta and Angiotensin II, disrupts critical cardiogenic regulators essential for heart function. Understanding these disruptions is crucial for developing targeted therapies and biomarkers to assess disease severity. This research addresses a crucial gap in cardiovascular treatment by focusing on mechanisms underlying remodeling processes, aiming to improve therapeutic strategies and outcomes for ischemic heart disease patients.

**Methods:** Databases, including PubMed, MEDLINE, Google Scholar, and open access/ subscription-based journals were searched for published articles without any date restrictions, to look into how TGF beta and Angiotensin II disrupt cardiogenic regulators in cardiac remodeling post-ischemic. Based on the criteria mentioned in the methods section, studies were systematically reviewed with focus on objectives of the study. This study adheres to relevant PRISMA guidelines (Preferred Reporting Items for Systematic Reviews and Meta-Analyses).

**Results:** Cardiac remodeling post-ischemic injury involves complex disruptions of cardiogenic regulators, prominently influenced by TGF-beta and Angiotensin II. Our study reveals these factors significantly alter critical regulators like Isl1, Nkx2–5, and GATA4, impacting myocardial repair mechanisms. TGF-beta induces fibrosis and inflammatory responses, while Angiotensin II exacerbates hypertrophic pathways and oxidative stress. Interactions between these pathways amplify remodeling through Smad, MAPK, and other signaling cascades. These findings point to the crucial roles of TGF-beta and Angiotensin II in pathological cardiac remodeling, highlighting potential targets for therapeutic interventions.

**Conclusion:** Cardiac remodeling post-ischemic injury, influenced by TGF-beta and Angiotensin II, disrupts vital cardiogenic regulators like Isl1, Brg1/Baf60 – Smarcd3 complex, Nkx2–5, GATA4, Tbx5, Mef2c, HAND1/2, MYOCD, MSX2, HOPX, Wnt-signaling pathway, Notch, FGF, and BMPs. These disruptions, involving altered receptor expression, signaling pathway interference, hypertrophic responses, and fibrosis promotion, compromise cardiac development and repair mechanisms. Targeting these pathways could enhance therapeutic strategies for ischemic heart disease by restoring normal regulator function and promoting effective cardiac repair and regeneration, thereby improving clinical outcomes.

## Background

Cardiac remodeling following ischemic injury represents a key challenge in cardiovascular medicine, characterized by structural and functional alterations that can lead to heart failure and adverse clinical outcomes. Ischemic events, such as myocardial infarction, initiate a cascade of molecular and cellular responses aimed at repairing damaged myocardium. However, prolonged or unresolved remodeling processes can exacerbate myocardial dysfunction and compromise overall cardiac function. Central to this process are signaling molecules like transforming growth factor-beta (TGF-beta) and Angiotensin II, which play critical roles in mediating fibrosis, hypertrophic responses, and inflammatory pathways within the myocardium post-ischemic injury [1]. These pathways have been implicated in disrupting a myriad of cardiogenic regulators essential for cardiac development and homeostasis. Understanding how TGF-beta and Angiotensin II disrupt cardiogenic regulators such as Isl1, Brg1/Baf60 – Smarcd3 complex, Nkx2–5, GATA4, Tbx5, Mef2c, HAND1/2, MYOCD, MSX2, HOPX, as well as Wnt-signaling pathway, Notch, FGF, and BMPs, is crucial for several reasons. It sheds light on the molecular mechanisms underlying cardiac remodeling post-ischemic injury, elucidating pathways that may be targeted for therapeutic intervention. Secondly, identifying specific disruptions in these regulatory networks provides potential biomarkers for assessing disease severity and predicting patient outcomes [2]. Moreover, this research addresses a significant gap in current cardiovascular therapies, where strategies primarily focus on symptom management rather than targeting the underlying molecular causes of cardiac dysfunction post-ischemic injury. By elucidating how TGF-beta and Angiotensin II dysregulate key cardiogenic regulators, this study aims to pave the way for novel therapeutic approaches aimed at mitigating adverse remodeling processes and improving long-term outcomes for patients with ischemic heart disease [3, 4].

## Methods

### Aim of the Study

The aim of this study is to investigate how cardiac remodeling occurs post-ischemic injury, with a particular focus on how TGF-β and Angiotensin II disrupt cardiogenic regulators. These regulators include Isl1, the Brg1/Baf60 (Smarcd3) complex, Nkx2-5, GATA4, Tbx5, Mef2c, HAND1/2, MYOCD, MSX2, HOPX, and pathways such as Wnt-signaling, Notch, FGF, and BMPs. The study seeks to look into the impact of these disruptions on the cardiogenic process and the resulting cardiac remodeling.

### Research Question

How do TGF-β and Angiotensin II disrupt cardiogenic regulators and pathways involved in cardiac remodeling post-ischemic injury?

### Search Focus

A comprehensive literature search was conducted using the PUBMED database, MEDLINE database, and Google Scholar, as well as open-access and subscription-based journals. There were no date restrictions for published articles. The search focused on TGF-β, Angiotensin II, and their effects on cardiogenic regulators and pathways, specifically Isl1, Brg1/Baf60 (Smarcd3) complex, Nkx2-5, GATA4, Tbx5, Mef2c, HAND1/2, MYOCD, MSX2, HOPX, Wnt-signaling pathway, Notch, FGF, and BMPs, in the context of cardiac remodeling post-ischemic injury.

### Cardiogenic Regulators and Pathways Investigated

#### Isl1, Brg1/Baf60 – Smarcd3 complex, Nkx2–5, GATA4, Tbx5, Mef2c, HAND1/2, MYOCD, MSX2, HOPX, Wnt-signaling pathway, Notch, FGF, BMPs

They were also used as search terms in search strategy in relation to TGF beta and Ag II, for their roles in cardiac remodeling. These factors and pathways were investigated to determine how TGF-β and Angiotensin II disrupt their normal function, leading to maladaptive cardiac remodeling post-ischemic injury. The literature search began in August 2020 and ended in November 2023. An in-depth investigation was conducted during this period based on the study parameters defined above. Further literature was searched and referenced until May 2024 during revision to maintain the highest accuracy. This comprehensive approach ensured that selected studies provided valuable insights into the disruptions caused by TGF-β and Angiotensin II in the cardiogenic process post-ischemic injury. This study adheres to relevant PRISMA guidelines (Preferred Reporting Items for Systematic Reviews and Meta-Analyses).

#### Screening and Eligibility Criteria

- **Initial Screening:** Articles were initially screened based on titles and abstracts to identify direct relevance to the study objectives.
- **Full-Text Review:** Articles that passed the initial screening underwent a comprehensive full-text review. Articles were included if they provided valuable insights into how TGF-β and Angiotensin II affect the specified cardiogenic regulators and pathways in cardiac remodeling post-ischemic injury.
- **Data Extraction:** Relevant data were extracted from each selected article, focusing on key findings and outcomes related to the study objectives.

#### Inclusion Criteria

- Articles directly related to TGF-β and Angiotensin II and their impact on the key cardiogenic regulators and pathways involved in cardiac remodeling post-ischemic injury.
- Studies focusing on the disruption of these factors and pathways by TGF-β and Angiotensin II.

#### Exclusion Criteria

- Articles that did not conform to the study focus.
- Insufficient methodological rigor.
- Data not aligning with the research questions.

### Rationale for Screening and Inclusion

The following transcription factors, genes, and signaling pathways were investigated based on their critical roles in the cardiogenic process and their implications in cardiac remodeling post-ischemic injury. Each factor has a well-established role in cardiogenesis and has been associated with disruptions in post-ischemic cardiac remodeling, making them pertinent to this study.

- **Isl1** is essential for cardiac progenitor cell development and differentiation. Disruption by TGF-β and Angiotensin II

can impair these processes, contributing to maladaptive remodeling.

- **Brg1/Baf60 (Smarcd3) complex** plays a crucial role in chromatin remodeling and gene expression in cardiogenesis. Its disruption can lead to altered gene expression and cardiac dysfunction.
- **Nkx2-5** is critical for cardiac development and function. TGF-β and Angiotensin II-mediated disruptions can lead to impaired cardiac remodeling.
- **GATA4** is involved in cardiac hypertrophy and development. Its dysregulation can result in pathological cardiac remodeling.
- **Tbx5** regulates heart development and is crucial for cardiac septation. Disruptions can lead to congenital heart defects and remodeling issues.
- **Mef2c** is essential for cardiac muscle development and function. TGF-β and Angiotensin II can alter its activity, leading to cardiac dysfunction.
- **HAND1/2** are involved in cardiac morphogenesis. Dysregulation can lead to defects in heart structure and function.
- **MYOCD** is a key regulator of cardiomyocyte differentiation and function. Disruption can impair heart muscle function.
- **MSX2** is involved in cardiac valve formation and development. Dysregulation can lead to valve defects and remodeling issues.
- **HOPX** regulates cardiomyocyte proliferation and differentiation. Its disruption can impair heart regeneration.
- **Wnt-signaling pathway** is critical for cardiac development and regeneration. Dysregulation can lead to cardiac fibrosis and impaired healing.
- **Notch** regulates cardiac progenitor cell fate. Disruption can lead to abnormal heart development and remodeling.
- **FGF** is involved in cardiac development and repair. Dysregulation can impair heart regeneration.
- **BMPs** regulate cardiac cell differentiation and function. Disruption can lead to congenital heart defects and remodeling issues.

This comprehensive screening and inclusion rationale ensure that the selected studies provide valuable insights into the roles of these critical factors in the cardiogenic process and their implications in cardiac remodeling post-ischemic injury.

### Exclusion Criteria

To ensure that only the most relevant and high-quality studies were included in this investigation, the following exclusion criteria were applied:

1. Non-Conformity with Study Focus: Articles that did not specifically focus on TGF-β, Angiotensin II, and their impact on Isl1, Brg1/Baf60 (Smarcd3) complex, Nkx2-5, GATA4, Tbx5, Mef2c, HAND1/2, MYOCD, MSX2, HOPX, Wnt-signaling pathway, Notch, FGF, and BMPs in cardiac remodeling post-ischemic injury.
2. Insufficient Methodological Rigor: Studies with poor experimental design, lack of appropriate controls, or inadequate statistical analysis.
3. Lack of Relevance to Post-Ischemic Cardiac Remodeling: Studies that did not explore the involvement of TGF-β and Angiotensin II in disrupting cardiogenic regulators and pathways post-ischemic injury.
4. Incomplete or Irrelevant Data: Articles that did not provide sufficient data or detailed analysis related to the key factors under investigation.
5. Duplicate Publications: Duplicate articles or those that presented redundant data from previously included studies.
6. Non-English Publications: Articles not published in the English language, to ensure consistency and comprehensibility of the reviewed literature.
7. Unpublished Studies: Unpublished studies, including conference abstracts and dissertations, were not included to ensure the inclusion of rigorously validated research.
8. Date of Publication: Although there were no date restrictions for the literature search, studies that did not align with current understanding and technological advancements were critically assessed for their relevance and methodological robustness.

By applying these exclusion criteria, the study maintained a high standard of evidence, ensuring that the included literature provided valuable and relevant insights into the roles of TGF-β and Angiotensin II in disrupting cardiogenic regulators and pathways in cardiac remodeling post-ischemic injury.

### Assessment of Article Quality and Potential Biases

Ensuring the quality and minimizing potential biases of the selected articles were crucial aspects to guarantee the rigor and reliability of the research findings.

- **Quality Assessment:** The initial step in quality assessment involved evaluating the methodological rigor of the selected articles. This included a thorough examination of the study design, data collection methods, and analyses conducted. The significance of the study’s findings was weighed based on the quality of the evidence presented. Articles demonstrating sound methodology—such as well-designed studies, controlled variables, and scientifically robust data—were considered of higher quality. Peer-reviewed articles, scrutinized by experts in the field, served as a significant indicator of quality.

#### • Potential Biases Assessment

`o` **Publication Bias:** To address the potential for publication bias, a comprehensive search strategy was adopted to include a balanced representation of both positive and negative results, incorporating a wide range of published articles from databases like Google Scholar.
`o` **Selection Bias:** Predefined and transparent inclusion criteria were applied to minimize subjectivity in the selection process. Articles were chosen based on their relevance to the study’s objectives, adhering strictly to these criteria. This approach reduced the risk of subjectivity and ensured that the selection process was objective and consistent.
`o` **Reporting Bias:** To mitigate reporting bias, articles were checked for inconsistencies or missing data. Multiple detailed reviews of the methodologies and results were conducted for all selected articles to identify and address any reporting bias.

By including high-quality, peer-reviewed studies and thoroughly assessing potential biases, this study aimed to provide a robust foundation for the results and conclusions presented.

### Language and Publication Restrictions

We restricted our selection to publications in the English language. There were no limitations imposed on the date of publication. Unpublished studies were not included in our analysis.

## Results

A total of 2680 articles were identified using database searching, and 2543 were recorded after duplicates removal. 2241 were excluded after screening of title/abstract, 180 were finally excluded, and 5 articles were excluded during data extraction. These exclusions were primarily due to factors such as non-conformity with the study focus, insufficient methodological rigor, or data that did not align with our research questions. Finally, 117 articles were included as references.

### Investigating Cardiogenic Mechanisms in Cardiac Remodeling

#### 1. Isl1

##### Effect of TGF-beta on this Cardiogenic Regulator

Islet1 (Isl1) is crucial in cardiac progenitor cell function during heart development, contributing to the formation of cardiac structures. In the context of cardiac repair, Isl1 promotes cardiomyocyte proliferation and differentiation, facilitating post-injury recovery [5]. TGF-β disrupts Isl1 expression in cardiomyocytes through multiple mechanisms. It activates SMAD2/3 signaling, leading to transcriptional repression and chromatin remodeling at the Isl1 promoter. Epigenetic modifications induced by TGF-β, such as DNA methylation and histone modifications, further suppress Isl1 expression, limiting cardiomyocyte regenerative potential. Additionally, TGF-β promotes cell cycle arrest and apoptosis in Isl1-positive cardiomyocytes by upregulating cyclin-dependent kinase inhibitors and activating pro-apoptotic pathways. This inhibition of proliferation and induction of cell death hinder cardiac regeneration post-ischemic injury. Moreover, TGF-β induces fibrosis by promoting myofibroblast differentiation and excessive extracellular matrix deposition [6]. The resulting fibrotic environment impedes Isl1-positive progenitor cell migration and function, exacerbating cardiac repair limitations. TGF-β also modulates critical signaling pathways like Wnt/β-catenin and Notch, crucial for Isl1 expression and cardiomyocyte proliferation. By antagonizing these pathways, TGF-β decreases Isl1 levels and disrupts regenerative processes. Furthermore, TGF-β induces inflammation and oxidative stress in the cardiac microenvironment, impairing Isl1-positive cell survival and differentiation. This chronic inflammatory response diminishes cardiac repair efficacy post-ischemic injury. Interactions with cardiogenic regulators like Nkx2-5, GATA4, and Tbx5 further inhibit Isl1 function through the formation of transcriptional repressive complexes [7]. Direct binding of TGF-β-induced transcriptional repressors to Isl1 promoters exacerbates these effects, hindering effective cardiac repair mechanisms. TGF-β exerts multidimensional effects on Isl1 in cardiomyocytes during cardiac remodeling post-ischemic injury, impairing regenerative pathways and promoting fibrosis, inflammation, and oxidative stress. Understanding these mechanisms informs potential therapeutic strategies to mitigate TGF-β’s adverse effects and enhance cardiac recovery [8].

##### Effect of Angiotensin II on this Cardiogenic Regulator

Islet1 (Isl1) serves as a critical transcription factor essential for the proliferation, differentiation, and survival of cardiac progenitor cells during heart development. These Isl1-positive progenitors contribute significantly to the formation of various cardiac structures, including the right ventricle, outflow tract, and parts of the atria. Moreover, Isl1 plays a key role in promoting the proliferation and differentiation of cardiomyocytes, thereby facilitating mechanisms involved in cardiac repair following injury [9]. Angiotensin II, a key player in cardiovascular physiology and pathology, exerts its influence on Isl1 in cardiomyocytes through several detrimental mechanisms. Angiotensin II binds to the angiotensin II type 1 receptor (AT1R), initiating downstream signaling pathways such as MAPK/ERK and JNK. Activation of these pathways can lead to the suppression of Isl1 expression by inducing transcriptional repressors or altering chromatin remodeling factors that inhibit Isl1 transcription. Additionally, Angiotensin II promotes cardiomyocyte hypertrophy via MAPK/ERK and PI3K/Akt signaling pathways. This hypertrophic response not only alters gene expression unfavorably, potentially diminishing the expression of regenerative genes like Isl1, but also shifts the cellular environment away from a regenerative state, reducing the pool of Isl1-positive cells available for cardiac repair [10]. Moreover, Angiotensin II stimulates fibroblast activation and differentiation into myofibroblasts, resulting in increased deposition of extracellular matrix (ECM) and subsequent fibrosis. This fibrotic environment hinders the migration and proliferation of Isl1-positive progenitor cells, further limiting their contribution to effective cardiac repair. The excessive ECM production and remodeling induced by Angiotensin II create a hostile microenvironment that compromises the survival and function of Isl1-positive cells. Furthermore, Angiotensin II triggers a pro-inflammatory response within the cardiac microenvironment, promoting the production of inflammatory cytokines (e.g., TNF-α, IL-6) and chemokines. Chronic inflammation induced by Angiotensin II adversely affects the survival and differentiation of Isl1-positive progenitor cells, thereby impairing cardiac repair mechanisms. Additionally, Angiotensin II-mediated inflammation increases oxidative stress levels, which can directly damage Isl1-positive cells and further diminish their regenerative capacity. Angiotensin II also interferes with key signaling pathways crucial for maintaining Isl1 expression and promoting cardiomyocyte proliferation, such as the Wnt/β-catenin and Notch pathways. By antagonizing these pathways, Angiotensin II reduces Isl1 levels and compromises the regenerative potential of cardiomyocytes. Furthermore, Angiotensin II induces apoptosis in cardiomyocytes through activation of pro-apoptotic signaling pathways like p53 and Bax, leading to decreased numbers of Isl1-positive cells available for cardiac repair. Moreover, Angiotensin II-induced cell cycle arrest in cardiomyocytes through upregulation of cyclin-dependent kinase inhibitors (e.g., p21, p27) further limits the proliferation of Isl1-positive cells essential for effective cardiac regeneration [11]. Angiotensin II disrupts Isl1 function through interactions with other cardiogenic regulators such as Nkx2-5, GATA4, and Tbx5, potentially forming repressive transcriptional complexes that inhibit Isl1 activity. These interactions and the binding of Angiotensin II-induced transcriptional repressors directly to the promoters of Isl1 and other cardiogenic genes contribute to their downregulation, exacerbating impaired cardiac repair processes. Angiotensin II exerts effects on Islet1 in cardiomyocytes during cardiac remodeling post-ischemic injury, including downregulation of Isl1 expression, promotion of hypertrophy and fibrosis, induction of pro-inflammatory effects, modulation of key signaling pathways, induction of apoptosis, and interaction with other cardiogenic regulators. Understanding these mechanisms provides a comprehensive insight into how Angiotensin II disrupts Isl1 function, thereby impairing cardiac repair and regeneration. This points to the potential for developing targeted therapeutic strategies aimed at mitigating the adverse effects of Angiotensin II and enhancing cardiac recovery following ischemic injury [12].

#### 2. Brg1/Baf60-Smarcd3 Complex

##### Effect of TGF-beta on this Cardiogenic Regulator

The Brg1/Baf60-Smarcd3 complex, a vital component of the SWI/SNF chromatin remodeling complex, is crucial for regulating gene expression by modifying chromatin structure. This complex plays a key role in cardiac development, influencing the differentiation, proliferation, and maintenance of cardiomyocytes essential for cardiac function. Brg1 (Brahma-related gene 1) and Baf60 (Smarcd3) are integral subunits that orchestrate the activation and repression of cardiac-specific genes crucial for heart development and repair [13]. TGF-β disrupts the Brg1/Baf60-Smarcd3 complex in cardiomyocytes through various mechanisms. TGF-β signaling can inhibit the chromatin remodeling activity of the complex by phosphorylating SMAD proteins. These phosphorylated SMADs interact with chromatin remodelers and transcription factors, potentially impairing the complex’s ability to regulate gene expression. Additionally, TGF-β induces epigenetic modifications such as DNA methylation and histone modifications, which can further hinder the recruitment and function of the Brg1/Baf60-Smarcd3 complex, thereby repressing genes essential for cardiac repair and regeneration. Moreover, TGF-β alters gene expression patterns by recruiting co-repressors and histone deacetylases (HDACs) to the promoters of target genes, leading to reduced transcriptional activity of the Brg1/Baf60-Smarcd3 complex [14]. This repression of cardiac-specific genes shifts the cellular environment towards fibrotic gene expression, contributing to adverse cardiac remodeling and increased fibrosis. The differentiation of fibroblasts into myofibroblasts induced by TGF-β further exacerbates ECM production, creating an environment hostile to cardiomyocytes and disrupting the function of the Brg1/Baf60-Smarcd3 complex. Furthermore, TGF-β interacts with key signaling pathways like Wnt/β-catenin, Notch, and Hippo pathways, which are crucial for cardiac development and repair. These interactions modulate the activity of the Brg1/Baf60-Smarcd3 complex, altering gene expression profiles essential for cardiomyocyte function and regeneration. TGF-β also induces oxidative stress and apoptosis in cardiomyocytes, leading to chromatin structural changes that impair the function of the Brg1/Baf60-Smarcd3 complex and hinder cardiac repair mechanisms. Additionally, TGF-β promotes a pro-inflammatory milieu by stimulating the production of cytokines and chemokines, creating chronic inflammation detrimental to cardiac repair. This persistent inflammation disrupts the function of the Brg1/Baf60-Smarcd3 complex by altering its ability to regulate gene expression and chromatin structure, further impairing cardiac recovery post-ischemic injury. Interactions with other cardiogenic regulators such as Nkx2-5, GATA4, and Tbx5 also contribute to the repression of gene expression by forming repressive transcriptional complexes that inhibit the activity of the Brg1/Baf60-Smarcd3 complex [15]. TGF-β exerts effects on the Brg1/Baf60-Smarcd3 complex in cardiomyocytes during cardiac remodeling post-ischemic injury, disrupting chromatin remodeling activity, altering gene expression, promoting fibrosis, modulating signaling pathways, inducing cellular stress and apoptosis, and interacting with other cardiogenic regulators. Understanding these mechanisms provides insights into how TGF-β impairs the function of the Brg1/Baf60-Smarcd3 complex, thereby compromising cardiac repair and regeneration [16].

##### Effect of Angiotensin II on this Cardiogenic Regulator

The Brg1/Baf60-Smarcd3 complex is integral to the SWI/SNF chromatin remodeling complex, crucial for regulating gene expression by modifying chromatin structure. This complex plays a key role in cardiac development, influencing the differentiation, proliferation, and maintenance of cardiomyocytes essential for cardiac function. Brg1 (Brahma-related gene 1) and Baf60 (Smarcd3) are key subunits that orchestrate the activation and repression of cardiac-specific genes critical for heart development and repair [17]. Angiotensin II disrupts the Brg1/Baf60-Smarcd3 complex in cardiomyocytes through several mechanisms. Through its receptor AT1R, Angiotensin II activates downstream signaling pathways such as MAPK/ERK, JNK, and NF-κB. These pathways can interact with chromatin remodelers and transcription factors, potentially inhibiting the chromatin remodeling activity of the Brg1/Baf60-Smarcd3 complex. Additionally, Angiotensin II induces epigenetic modifications like DNA methylation and histone modifications, altering the recruitment and activity of the complex, and leading to repression of genes crucial for cardiac repair and regeneration. The Brg1/Baf60-Smarcd3 complex is vital for activating cardiac-specific genes, and Angiotensin II can repress these genes by recruiting co-repressors and histone deacetylases (HDACs) to their promoters. This repression shifts cellular gene expression towards hypertrophic and fibrotic pathways, contributing to adverse cardiac remodeling [18]. Moreover, Angiotensin II induces cardiomyocyte hypertrophy through pathways like MAPK/ERK and PI3K/Akt, altering gene expression that may negatively impact regenerative genes controlled by the Brg1/Baf60-Smarcd3 complex. Furthermore, Angiotensin II promotes fibroblast activation and differentiation into myofibroblasts, leading to increased ECM deposition and fibrosis. This excessive ECM production creates an environment hostile to cardiomyocytes and disrupts the Brg1/Baf60-Smarcd3 complex’s ability to regulate genes essential for cardiac repair. Angiotensin II also stimulates pro-inflammatory cytokine production and oxidative stress in the cardiac microenvironment, further impairing the complex’s function and hindering effective cardiac repair. Angiotensin II interacts with pathways crucial for cardiac development and repair, such as Wnt/β-catenin, Notch, and Hippo pathways, modulating the Brg1/Baf60-Smarcd3 complex’s activity and altering gene expression profiles in cardiomyocytes [19]. Additionally, Angiotensin II-induced cellular stress and apoptosis in cardiomyocytes can lead to chromatin structural changes that impair the complex’s function and disrupt cardiac repair mechanisms. Moreover, interactions with cardiogenic regulators like Nkx2-5, GATA4, and Tbx5 contribute to the formation of repressive transcriptional complexes that inhibit the Brg1/Baf60-Smarcd3 complex’s activity. This transcriptional repression directly impacts genes crucial for cardiac repair processes. Angiotensin II impacts the Brg1/Baf60-Smarcd3 complex in cardiomyocytes during cardiac remodeling post-ischemic injury through multiple pathways, including altering chromatin remodeling activity, regulating gene expression, promoting hypertrophy and fibrosis, inducing pro-inflammatory effects, modulating key signaling pathways, inducing cellular stress and apoptosis, and interacting with other cardiogenic regulators. Understanding these mechanisms provides a comprehensive perspective on how Angiotensin II disrupts the Brg1/Baf60-Smarcd3 complex, thereby impairing cardiac repair and regeneration. This knowledge can guide the development of targeted therapies to mitigate Angiotensin II’s adverse effects and enhance cardiac recovery following ischemic injury [20].

#### 3. Nkx2–5

##### Effect of TGF-beta on this Cardiogenic Regulator

Nkx2-5, a crucial transcription factor in cardiac development and function, plays essential roles in cardiomyocyte differentiation, proliferation, survival, and the regulation of genes crucial for cardiac structure and function throughout various developmental stages and in adult cardiac homeostasis [21]. Its involvement spans from early heart tube formation to the maturation of cardiac chambers, demonstrating its significance in maintaining cardiac integrity and facilitating repair mechanisms in the adult heart. TGF-β exerts diverse effects on Nkx2-5 in cardiomyocytes during cardiac remodeling following ischemic injury. TGF-β signaling can lead to the downregulation of Nkx2-5 expression through transcriptional repression mechanisms. This involves the recruitment of transcriptional repressors and chromatin modifiers to the Nkx2-5 promoter, resulting in suppressed Nkx2-5 gene transcription and perturbing its critical regulatory network. Additionally, TGF-β induces epigenetic modifications such as DNA methylation and histone modifications, which contribute to a more repressive chromatin environment at the Nkx2-5 locus, further diminishing its expression levels [22]. Moreover, TGF-β affects Nkx2-5 protein stability through post-translational modifications like phosphorylation, ubiquitination, or acetylation. These modifications can influence Nkx2-5 stability, localization, and functional activity, potentially leading to protein degradation or functional inactivation. TGF-β signaling also involves cross-talk with Smad proteins, which interact with Nkx2-5 to modulate its transcriptional activity. Depending on the context and presence of co-factors, these interactions can either enhance or repress Nkx2-5-dependent transcriptional programs, impacting its role in cardiac repair. Furthermore, TGF-β promotes fibrosis by stimulating the differentiation of fibroblasts into myofibroblasts, resulting in increased extracellular matrix deposition and cardiac stiffening. This altered cardiac environment compromises Nkx2-5 function and its ability to regulate target genes essential for cardiomyocyte integrity and function [23]. Additionally, TGF-β-induced inflammation exacerbates oxidative stress and production of pro-inflammatory cytokines, which further impairs Nkx2-5 expression and function, hindering effective cardiac repair processes. TGF-β-induced cellular stress and apoptosis in cardiomyocytes contribute to additional disruptions in Nkx2-5 function. Oxidative stress damages cardiomyocytes and disrupts Nkx2-5 activity, affecting chromatin structure and gene expression patterns crucial for cardiac repair mechanisms. Moreover, TGF-β can induce apoptosis through pro-apoptotic signaling pathways in cardiomyocytes, leading to cardiomyocyte loss and exacerbating inflammatory responses that adversely affect Nkx2-5 function. Finally, TGF-β interacts with other cardiogenic regulators such as GATA4, Tbx5, and Mef2c, forming repressive transcriptional complexes that inhibit Nkx2-5 activity. These interactions involve TGF-β-induced transcriptional repressors directly binding to promoters of genes regulated by Nkx2-5, further impairing their expression and disrupting cardiac repair processes. TGF-β impacts Nkx2-5 in cardiomyocytes during cardiac remodeling post-ischemic injury through a multitude of mechanisms, encompassing transcriptional repression, protein stability alteration, modulation of signaling pathways, promotion of fibrosis, pro-inflammatory effects, induction of cellular stress and apoptosis, and interaction with other cardiogenic regulators. Understanding these mechanisms provides comprehensive insights into how TGF-β disrupts Nkx2-5 function, thereby impairing cardiac repair and regeneration. This knowledge holds promise for informing targeted therapeutic strategies aimed at mitigating the adverse effects of TGF-β and enhancing cardiac recovery following ischemic injury [24].

##### Effect of Angiotensin II on this Cardiogenic Regulator

Nkx2-5, a critical transcription factor in cardiac development, plays crucial roles in regulating cardiomyocyte differentiation, proliferation, and survival. It is essential for maintaining cardiac structure and function throughout various stages of heart development, from early formation to the maturation of cardiac chambers. In the adult heart, Nkx2-5 also contributes significantly to cardiac homeostasis and repair mechanisms [25]. Angiotensin II impacts Nkx2-5 in cardiomyocytes during cardiac remodeling post-ischemic injury through several mechanisms. Angiotensin II activates signaling pathways such as NF-κB, JNK, and MAPK/ERK, which can lead to transcriptional repression of Nkx2-5. This involves the recruitment of transcriptional repressors and chromatin modifiers to the Nkx2-5 promoter, suppressing its gene expression and disrupting the transcriptional network critical for cardiomyocyte function and survival. Additionally, Angiotensin II induces epigenetic modifications like DNA methylation and histone modifications, which further repress Nkx2-5 transcription by creating a more repressive chromatin environment. Moreover, Angiotensin II influences Nkx2-5 protein stability through post-translational modifications such as phosphorylation, ubiquitination, or acetylation. These modifications can affect the stability, localization, and activity of Nkx2-5, potentially leading to its degradation or functional inactivation [26]. Angiotensin II signaling also involves cross-talk with other pathways like MAPK/ERK, NF-κB, and PI3K/Akt, which can either enhance or repress Nkx2-5-dependent transcriptional programs depending on context and co-factor presence. Furthermore, Angiotensin II inhibits pro-regenerative pathways crucial for cardiac repair, including the Wnt/β-catenin pathway, indirectly affecting Nkx2-5 function and its role in cardiac repair processes. Angiotensin II promotes cardiomyocyte hypertrophy through pathways like MAPK/ERK and PI3K/Akt, shifting the cellular environment away from a regenerative state and potentially reducing the expression of Nkx2-5-regulated genes essential for cardiac repair. Additionally, Angiotensin II stimulates fibroblast activation and differentiation into myofibroblasts, leading to increased extracellular matrix deposition and fibrosis. This fibrotic environment can impair Nkx2-5 function and disrupt the transcriptional regulation of its target genes, affecting cardiomyocyte integrity and function. Furthermore, Angiotensin II induces pro-inflammatory effects by stimulating the production of inflammatory cytokines and chemokines, exacerbating oxidative stress, and damaging cardiomyocytes. This inflammatory response negatively impacts Nkx2-5 expression and function, hindering effective cardiac repair mechanisms. Angiotensin II-induced cellular stress and apoptosis in cardiomyocytes further disrupt Nkx2-5 function, altering chromatin structure and gene expression patterns crucial for cardiac repair. Finally, Angiotensin II interacts with other cardiogenic regulators such as GATA4, Tbx5, and Mef2c, forming repressive transcriptional complexes that inhibit Nkx2-5 activity. These interactions involve Angiotensin II-induced transcriptional repressors directly binding to promoters of genes regulated by Nkx2-5, further impairing their expression and disrupting cardiac repair processes. Angiotensin II impacts Nkx2-5 in cardiomyocytes during cardiac remodeling post-ischemic injury through a complex array of mechanisms, including downregulation of Nkx2-5 expression, alteration of Nkx2-5 protein stability, modulation of signaling pathways, promotion of hypertrophy and fibrosis, pro-inflammatory effects, induction of cellular stress and apoptosis, and interaction with other cardiogenic regulators [27]. Understanding these mechanisms provides a comprehensive perspective on how Angiotensin II disrupts Nkx2-5 function, thereby impairing cardiac repair and regeneration. This understanding could pave the way for developing targeted therapeutic strategies aimed at mitigating the adverse effects of Angiotensin II and enhancing cardiac recovery following ischemic injury [28].

#### 4. GATA4

##### Effect of TGF-beta on this Cardiogenic Regulator

GATA4, a critical transcription factor in cardiac development, plays essential roles in regulating cardiomyocyte differentiation, survival, and adaptive responses to stress. It is crucial in cardiac morphogenesis, hypertrophy, apoptosis, and the integration of mechanical and hormonal signals throughout heart development and adult cardiac function. TGF-β exerts significant impacts on GATA4 in cardiomyocytes during cardiac remodeling following ischemic injury through diverse mechanisms [29]. TGF-β signaling activates Smad proteins, which interact with transcriptional repressors and chromatin modifiers, thereby repressing the GATA4 promoter. This repression reduces GATA4 gene expression, disrupting its regulatory network within cardiomyocytes. Additionally, TGF-β induces epigenetic modifications like DNA methylation and histone modifications that further suppress GATA4 transcription, establishing a repressive chromatin landscape at the GATA4 locus. Furthermore, TGF-β influences GATA4 protein stability through post-translational modifications such as phosphorylation, ubiquitination, or acetylation. These modifications impact GATA4’s stability, localization, and activity, potentially leading to its degradation or functional inactivation. TGF-β also modulates signaling pathways crucial for cardiac regeneration, such as the Wnt/β-catenin pathway, which indirectly affects GATA4 function and its role in cardiac repair. Moreover, TGF-β promotes fibrosis by stimulating fibroblast differentiation into myofibroblasts, resulting in increased extracellular matrix deposition and cardiac stiffness [30]. This altered ECM environment compromises GATA4 function and disrupts the transcriptional regulation of its target genes, crucial for maintaining cardiomyocyte integrity. Additionally, TGF-β-induced inflammation triggers the production of pro-inflammatory cytokines and chemokines, exacerbating oxidative stress and damaging cardiomyocytes, thereby negatively impacting GATA4 expression and function. TGF-β-induced cellular stress and apoptosis further contribute to GATA4 dysfunction by altering chromatin structure and gene expression patterns necessary for cardiac repair mechanisms. Additionally, TGF-β interacts with other cardiogenic regulators such as Nkx2-5, Tbx5, and Mef2c, forming repressive transcriptional complexes that inhibit GATA4 activity [31]. These interactions involve TGF-β-induced transcriptional repressors directly binding to promoters of genes regulated by GATA4, further impairing their expression and disrupting cardiac repair processes. TGF-β disrupts GATA4 function in cardiomyocytes during cardiac remodeling post-ischemic injury through a complex array of mechanisms, including downregulation of GATA4 expression, alteration of GATA4 protein stability, modulation of signaling pathways, promotion of fibrosis, pro-inflammatory effects, induction of cellular stress and apoptosis, and interaction with other cardiogenic regulators. Understanding these mechanisms provides a comprehensive insight into how TGF-β hinders GATA4-mediated cardiac repair and regeneration. This knowledge could facilitate the development of targeted therapeutic strategies aimed at mitigating the adverse effects of TGF-β and enhancing cardiac recovery following ischemic injury [32].

##### Effect of Angiotensin II on this Cardiogenic Regulator

GATA4, a critical transcription factor in cardiac development, plays crucial roles in regulating cardiomyocyte differentiation, proliferation, survival, and adaptive responses to stress. It is essential for cardiac morphogenesis, myocardial survival, and responses to mechanical and hormonal signals throughout cardiac development and adulthood. Angiotensin II exerts significant impacts on GATA4 in cardiomyocytes during cardiac remodeling following ischemic injury through diverse mechanisms [33]. Angiotensin II activates signaling pathways such as NF-κB, JNK, and MAPK/ERK, which recruit transcriptional repressors and chromatin modifiers to the GATA4 promoter, suppressing GATA4 gene expression and impairing its regulatory network in cardiomyocytes. Additionally, Angiotensin II induces epigenetic modifications like DNA methylation and histone modifications, creating a repressive chromatin environment at the GATA4 locus, further reducing its expression. Furthermore, Angiotensin II alters GATA4 protein stability through post-translational modifications such as phosphorylation, ubiquitination, or acetylation. These modifications influence GATA4 stability, localization, and activity, potentially leading to its degradation or functional inactivation. Angiotensin II also modulates signaling pathways crucial for cardiac regeneration, including MAPK/ERK, NF-κB, and PI3K/Akt, which can either enhance or suppress GATA4-dependent transcriptional programs depending on context and co-factor presence. Inhibition of pathways essential for cardiac regeneration, such as the Wnt/β-catenin pathway, indirectly affects GATA4 function and its role in cardiac repair [34]. Moreover, Angiotensin II promotes cardiomyocyte hypertrophy through MAPK/ERK and PI3K/Akt signaling pathways, shifting the cellular environment away from a regenerative state and reducing the pool of GATA4-regulated genes essential for cardiac repair. This hypertrophic response involves alterations in gene expression that may negatively impact the expression of regenerative genes regulated by GATA4. Additionally, Angiotensin II promotes fibroblast activation and differentiation into myofibroblasts, increasing extracellular matrix deposition and causing fibrosis. This fibrotic tissue alters the cardiac environment, impairing GATA4 function and disrupting the transcriptional regulation of its target genes.

Excessive extracellular matrix production and remodeling physically disrupt cell-cell and cell-ECM interactions crucial for GATA4 function and cardiomyocyte integrity. Furthermore, Angiotensin II stimulates the production of pro-inflammatory cytokines (e.g., TNF-α, IL-6) and chemokines, triggering chronic inflammation that negatively impacts GATA4 expression and function. This inflammatory response also leads to increased oxidative stress, damaging cardiomyocytes and further disrupting GATA4 activity. Angiotensin II-induced cellular stress and apoptosis contribute to GATA4 dysfunction by altering chromatin structure and gene expression patterns necessary for effective cardiac repair mechanisms [35]. Additionally, Angiotensin II interacts with other cardiogenic regulators such as Nkx2-5, Tbx5, and Mef2c, forming repressive transcriptional complexes that inhibit GATA4 activity. These interactions involve Angiotensin II-induced transcriptional repressors directly binding to promoters of genes regulated by GATA4, impairing their expression and disrupting cardiac repair processes. Angiotensin II disrupts GATA4 function in cardiomyocytes during cardiac remodeling post-ischemic injury through multiple mechanisms, including downregulation of GATA4 expression, alteration of GATA4 protein stability, modulation of signaling pathways, promotion of hypertrophy and fibrosis, pro-inflammatory effects, induction of cellular stress and apoptosis, and interaction with other cardiogenic regulators. Understanding these mechanisms provides a comprehensive view of how Angiotensin II impairs GATA4-mediated cardiac repair and regeneration. This knowledge can guide the development of targeted therapeutic strategies aimed at mitigating the adverse effects of Angiotensin II and enhancing cardiac recovery following ischemic injury [36].

#### 5. Tbx5

##### Effect of TGF-beta on this Cardiogenic Regulator

Tbx5, a critical transcription factor in cardiac development, is integral to the differentiation, proliferation, and survival of cardiomyocytes, as well as the regulation of genes essential for cardiac morphogenesis and conduction system development. It plays a significant role in maintaining cardiac homeostasis and repair mechanisms throughout both cardiac development and in the adult heart. TGF-β exerts profound effects on Tbx5 in cardiomyocytes during cardiac remodeling following ischemic injury through various mechanisms. TGF-β signaling activates Smad proteins, which interact with transcriptional repressors and chromatin modifiers to suppress the Tbx5 promoter, resulting in decreased Tbx5 gene expression and impairing its regulatory network in cardiomyocytes [37]. Additionally, TGF-β induces epigenetic modifications like DNA methylation and histone modifications, establishing a repressive chromatin environment at the Tbx5 locus that reduces its expression. Moreover, TGF-β influences Tbx5 protein stability through post-translational modifications such as phosphorylation, ubiquitination, or acetylation, which can affect Tbx5 stability, localization, and activity, potentially leading to its degradation or functional inactivation. TGF-β also modulates signaling pathways crucial for cardiac regeneration, including interactions with Smad proteins that can either enhance or suppress Tbx5-dependent transcriptional programs depending on cellular context and co-factor presence. Inhibition of pathways essential for cardiac regeneration, such as the Wnt/β-catenin pathway, indirectly impacts Tbx5 function and its role in cardiac repair [38]. Furthermore, TGF-β promotes fibroblast activation and differentiation into myofibroblasts, leading to increased extracellular matrix (ECM) deposition and fibrosis. This fibrotic environment alters the cardiac milieu, impairing Tbx5 function and disrupting the transcriptional regulation of its target genes. Excessive ECM production and remodeling also physically disrupt cell-cell and cell-ECM interactions crucial for Tbx5 function and cardiomyocyte integrity. Additionally, TGF-β stimulates the production of pro-inflammatory cytokines (e.g., TNF-α, IL-6) and chemokines, contributing to chronic inflammation that negatively impacts Tbx5 expression and function, thereby hindering effective cardiac repair. TGF-β-induced oxidative stress further exacerbates these effects by damaging cardiomyocytes and disrupting Tbx5 activity. Moreover, TGF-β-induced cellular stress and apoptosis in cardiomyocytes activate pro-apoptotic signaling pathways, leading to cardiomyocyte loss and an inflammatory response that further disrupts Tbx5 activity. TGF-β’s interactions with other cardiogenic regulators, such as Nkx2-5, GATA4, and Mef2c, also play a role in forming repressive transcriptional complexes that inhibit Tbx5 activity. These interactions include TGF-β-induced transcriptional repressors directly binding to promoters of genes regulated by Tbx5, leading to their downregulation and impairing cardiac repair processes [39].

TGF-β disrupts Tbx5 function in cardiomyocytes during cardiac remodeling post-ischemic injury through multiple mechanisms, including downregulation of Tbx5 expression, alteration of Tbx5 protein stability, modulation of signaling pathways, promotion of fibrosis, pro-inflammatory effects, induction of cellular stress and apoptosis, and interaction with other cardiogenic regulators. Understanding these mechanisms provides a comprehensive perspective on how TGF-β impairs Tbx5-mediated cardiac repair and regeneration. This knowledge is crucial for developing targeted therapeutic strategies aimed at mitigating the adverse effects of TGF-β and enhancing cardiac recovery following ischemic injury [40].

##### Effect of Angiotensin II on this Cardiogenic Regulator

Angiotensin II exerts significant impacts on Tbx5 in cardiomyocytes during cardiac remodeling post-ischemic injury through various mechanisms. Angiotensin II activates signaling pathways such as NF-κB, JNK, and MAPK/ERK, which recruit transcriptional repressors and chromatin modifiers to the Tbx5 promoter. This suppresses Tbx5 gene expression, thereby impairing its regulatory network in cardiomyocytes. Additionally, Angiotensin II induces epigenetic modifications like DNA methylation and histone modifications, establishing a repressive chromatin environment at the Tbx5 locus and reducing its expression [41]. Moreover, Angiotensin II influences Tbx5 protein stability via post-translational modifications such as phosphorylation, ubiquitination, or acetylation, which can alter Tbx5 stability, localization, and activity, potentially leading to its degradation or functional inactivation. Angiotensin II also modulates signaling pathways critical for cardiac regeneration, including interactions with pathways like MAPK/ERK, NF-κB, and PI3K/Akt, which can either enhance or suppress Tbx5-dependent transcriptional programs based on cellular context and co-factor presence. Inhibition of pathways essential for cardiac regeneration, such as the Wnt/β-catenin pathway, indirectly impacts Tbx5 function and its role in cardiac repair. Furthermore, Angiotensin II is an important inducer of cardiomyocyte hypertrophy through signaling pathways like MAPK/ERK and PI3K/Akt, shifting the cellular environment away from a regenerative state and reducing the pool of Tbx5-regulated genes [42].

This hypertrophic response involves changes in gene expression that may adversely impact the expression of regenerative genes regulated by Tbx5. Additionally, Angiotensin II promotes fibroblast activation and differentiation into myofibroblasts, leading to increased extracellular matrix (ECM) deposition and fibrosis. The resultant fibrotic tissue alters the cardiac environment, impairing Tbx5 function and disrupting the transcriptional regulation of its target genes. Excessive ECM production and remodeling also physically disrupt cell-cell and cell-ECM interactions crucial for Tbx5 function and cardiomyocyte integrity. Moreover, Angiotensin II stimulates pro-inflammatory cytokine (e.g., TNF-α, IL-6) and chemokine production, contributing to chronic inflammation that negatively affects Tbx5 expression and function, thereby hindering effective cardiac repair [43]. Angiotensin II-induced oxidative stress further exacerbates these effects by damaging cardiomyocytes and disrupting Tbx5 activity. Furthermore, Angiotensin II-induced cellular stress and apoptosis in cardiomyocytes activate pro-apoptotic signaling pathways, leading to cardiomyocyte loss and an inflammatory response that further disrupts Tbx5 activity. Angiotensin II’s interactions with other cardiogenic regulators such as Nkx2-5, GATA4, and Mef2c can form repressive transcriptional complexes that inhibit Tbx5 activity. This includes Angiotensin II-induced transcriptional repressors directly binding to promoters of genes regulated by Tbx5, leading to their downregulation and impairing cardiac repair processes [44].

#### 6. Mef2c

##### Effect of TGF-beta on this Cardiogenic Regulator

Mef2c is a critical transcription factor crucial in cardiac development, influencing the differentiation, proliferation, and survival of cardiomyocytes. It orchestrates the regulation of genes crucial for cardiac morphogenesis, including structural proteins and signaling molecules essential for heart formation and function. Additionally, Mef2c contributes to adaptive responses to stress and plays a significant role in cardiac remodeling and repair mechanisms within the adult heart [45]. TGF-β exerts impact on Mef2c in cardiomyocytes during cardiac remodeling post-ischemic injury through several mechanisms. TGF-β signaling activates Smad proteins, which can collaborate with transcriptional repressors and chromatin modifiers to suppress the Mef2c promoter, thereby diminishing Mef2c gene expression. Concurrently, TGF-β induces epigenetic modifications like DNA methylation and histone modifications, establishing a repressive chromatin environment at the Mef2c locus and further reducing its expression. Moreover, TGF-β influences Mef2c protein stability via post-translational modifications such as phosphorylation, ubiquitination, or acetylation, which can alter Mef2c stability, localization, and activity, potentially leading to its degradation or functional inactivation [46]. TGF-β also modulates signaling pathways crucial for cardiac regeneration, including interactions with Smad proteins and components of pathways like Wnt/β-catenin, which indirectly impact Mef2c function and its role in cardiac repair. Additionally, TGF-β promotes fibroblast activation and differentiation into myofibroblasts, contributing to increased deposition of extracellular matrix (ECM) and fibrosis. This fibrotic tissue alters the cardiac environment, impairing Mef2c function and disrupting the transcriptional regulation of its target genes. Excessive ECM production and remodeling further physically disrupt cell-cell and cell-ECM interactions crucial for Mef2c function and maintaining cardiomyocyte integrity. Furthermore, TGF-β stimulates the production of pro-inflammatory cytokines and chemokines, fostering chronic inflammation that negatively affects Mef2c expression and function, thereby hindering effective cardiac repair. Additionally, TGF-β-induced oxidative stress exacerbates these effects by damaging cardiomyocytes and disrupting Mef2c activity. Moreover, TGF-β-induced cellular stress and apoptosis in cardiomyocytes activate pro-apoptotic signaling pathways, resulting in cardiomyocyte loss and an inflammatory response that further disrupts Mef2c activity. TGF-β also interacts with other cardiogenic regulators such as Nkx2-5, GATA4, and Tbx5, potentially forming repressive transcriptional complexes that inhibit Mef2c activity [47]. This includes TGF-β-induced transcriptional repressors directly binding to promoters of genes regulated by Mef2c, leading to their downregulation and impairing processes critical for cardiac repair. TGF-β impacts Mef2c in cardiomyocytes during cardiac remodeling post-ischemic injury through a spectrum of mechanisms, encompassing downregulation of Mef2c expression, alteration of Mef2c protein stability, modulation of signaling pathways, promotion of fibrosis, pro-inflammatory effects, induction of cellular stress and apoptosis, and interaction with other cardiogenic regulators. Understanding these mechanisms provides a comprehensive perspective on how TGF-β disrupts Mef2c-mediated cardiac repair and regeneration. This knowledge is crucial for developing targeted therapeutic strategies aimed at mitigating the adverse effects of TGF-β and enhancing cardiac recovery following ischemic injury [48].

##### Effect of Angiotensin II on this Cardiogenic Regulator

Angiotensin II exerts profound effects on Mef2c in cardiomyocytes, impacting cardiac remodeling post-ischemic injury through several interconnected pathways. Angiotensin II activates signaling cascades such as NF-κB and MAPK/ERK, which recruit transcriptional repressors to the Mef2c promoter [49]. This transcriptional repression reduces Mef2c gene expression, disrupting its critical role in regulating cardiomyocyte function and repair processes. Furthermore, Angiotensin II inhibits pro-regenerative pathways essential for cardiac repair, including the Wnt/β-catenin pathway that interacts closely with Mef2c. This inhibition indirectly hampers Mef2c function, impairing its ability to promote effective cardiac repair mechanisms in response to ischemic injury. Angiotensin II-induced oxidative stress in cardiomyocytes contributes significantly to Mef2c dysfunction by causing cellular damage and functional impairment. This oxidative environment alters Mef2c activity and stability, diminishing its capacity to regulate gene expression crucial for cardiac adaptation and repair. Concurrently, Angiotensin II promotes a pro-inflammatory milieu by stimulating cytokine production and immune cell activation. Chronic inflammation adversely affects Mef2c expression and function, exacerbating the impairment of cardiac recovery post-ischemic injury [50]. Moreover, Angiotensin II activates hypertrophic signaling pathways like MAPK/ERK and PI3K/Akt in cardiomyocytes, promoting cardiomyocyte hypertrophy. This hypertrophic state may divert resources away from regenerative processes governed by Mef2c, further hindering cardiac repair mechanisms. Additionally, Angiotensin II induces fibroblast activation and differentiation into myofibroblasts, resulting in increased extracellular matrix (ECM) deposition and fibrosis within the cardiac tissue. The stiffened ECM disrupts essential cellular interactions necessary for Mef2c function, thereby compromising its regulatory role in cardiomyocyte function and repair. Furthermore, Angiotensin II may interact synergistically with other cardiogenic regulators such as GATA4 and Nkx2-5, forming repressive transcriptional complexes that inhibit Mef2c activity [51]. These interactions exacerbate the impairment of Mef2c-mediated cardiac repair mechanisms following ischemic injury. Angiotensin II disrupts Mef2c function in cardiomyocytes during cardiac remodeling post-ischemic injury through multiple intertwined mechanisms. These include transcriptional repression, modulation of signaling pathways, induction of oxidative stress and inflammation, promotion of cardiomyocyte hypertrophy, ECM remodeling, and interaction with other cardiogenic regulators. Understanding these mechanisms provides crucial insights into how Angiotensin II contributes to impaired cardiac repair and regeneration, suggesting potential therapeutic avenues to enhance Mef2c-mediated recovery in ischemic heart disease [52].

#### 7. HAND1/2

##### Effect of TGF-beta on this Cardiogenic Regulator

TGF-β exerts significant effects on HAND1 and HAND2 in cardiomyocytes during cardiac remodeling post-ischemic injury through various complex mechanisms. HAND1 and HAND2 are critical transcription factors essential for cardiac development and function. They regulate gene expression patterns that govern cell proliferation, differentiation, and tissue patterning during embryonic heart development [53]. Even in adult cardiomyocytes, HAND proteins continue to play crucial roles in cardiac remodeling and adaptation in response to stress or injury. One of the primary mechanisms by which TGF-β disrupts HAND1/2 in cardiomyocytes is through transcriptional repression. TGF-β signaling activates Smad proteins, which directly interact with the promoters of HAND1/2 genes, leading to the repression of their transcription. This reduction in HAND1/2 expression diminishes their regulatory functions in controlling cardiac remodeling and repair processes. Additionally, TGF-β modulates signaling pathways that cross-talk with Smad proteins and other pathways involved in cardiac development, such as BMP signaling. These interactions can synergistically inhibit HAND1/2 expression or activity through complex regulatory mechanisms, further impairing their roles in maintaining cardiac homeostasis and adaptive responses. Moreover, TGF-β is a key inducer of fibrosis within the heart, promoting the differentiation of fibroblasts into myofibroblasts and increasing extracellular matrix (ECM) deposition [54]. The fibrotic environment alters cardiomyocyte interactions and signaling, indirectly impacting HAND1/2 function and their regulatory networks critical for cardiac repair. Furthermore, TGF-β stimulates the production of pro-inflammatory cytokines and chemokines, contributing to chronic inflammation in the heart. This inflammatory milieu disrupts HAND1/2 expression and function, impairing their ability to regulate cardiomyocyte gene expression and participate effectively in cardiac remodeling processes. Additionally, TGF-β signaling pathways include components that promote apoptosis in cardiomyocytes under conditions of stress or injury. Increased apoptosis can lead to reduced HAND1/2 expression and function, diminishing their roles in promoting cardiac repair and remodeling. TGF-β may interact synergistically with other transcription factors or regulatory proteins involved in cardiomyocyte development and function, such as GATA4 or Mef2c. These interactions can form inhibitory complexes that suppress HAND1/2 activity or expression levels, further compromising their function in facilitating cardiac repair processes. TGF-β disrupts HAND1/2 function in cardiomyocytes during cardiac remodeling post-ischemic injury through multiple interconnected mechanisms. These include transcriptional repression mediated by Smad proteins, modulation of signaling pathways, induction of fibrosis and inflammation, promotion of apoptosis, and interactions with other cardiogenic regulators [55, 56].

##### Effect of Angiotensin II on this Cardiogenic Regulator

Angiotensin II significantly impacts HAND1 and HAND2 in cardiomyocytes during cardiac remodeling post-ischemic injury through various complex mechanisms. HAND1 and HAND2 are crucial transcription factors essential for cardiac function, regulating gene expression patterns that control cardiomyocyte function and repair processes. One primary effect of Angiotensin II is the modulation of transcriptional activity. It activates pathways such as NF-κB and MAPK/ERK, leading to the recruitment of transcriptional repressors to the HAND1/2 gene promoters. This repression reduces HAND1/2 gene expression, disrupting their regulatory role in maintaining cardiomyocyte function and impairing repair mechanisms [57]. Additionally, Angiotensin II interacts with the Renin-Angiotensin System (RAS), a key regulator of cardiac homeostasis and remodeling. This interaction modulates intracellular signaling pathways that intersect with HAND1/2-regulated pathways, potentially altering their transcriptional activity and functional outcomes in cardiomyocytes. Moreover, Angiotensin II induces a hypertrophic response in cardiomyocytes by activating pathways like MAPK/ERK and PI3K/Akt, promoting hypertrophic growth. This shift in cellular state may divert resources away from regenerative processes regulated by HAND1/2, thereby hindering effective cardiac repair.

Furthermore, Angiotensin II stimulates fibroblast activation, leading to differentiation into myofibroblasts and increased deposition of extracellular matrix (ECM), resulting in fibrosis. The stiffened ECM physically disrupts cellular interactions crucial for HAND1/2 function, impacting their regulatory role in cardiomyocyte physiology and repair processes [58]. Angiotensin II also promotes a pro-inflammatory environment by inducing cytokine production and immune cell activation. Chronic inflammation negatively affects HAND1/2 expression and function, further impairing cardiac recovery following ischemic injury. Additionally, Angiotensin II induces oxidative stress in cardiomyocytes, generating reactive oxygen species (ROS) that cause damage and functional impairment. This oxidative environment alters the activity and stability of HAND1/2, compromising their ability to regulate gene expression critical for cardiac adaptation and repair. Finally, Angiotensin II interacts synergistically with other cardiogenic regulators such as GATA4 or Mef2c, forming inhibitory complexes that suppress HAND1/2 activity or expression levels. These interactions further diminish HAND1/2-mediated cardiac repair mechanisms in response to ischemic injury. Angiotensin II disrupts HAND1/2 function in cardiomyocytes during cardiac remodeling post-ischemic injury through multiple interconnected mechanisms. These include transcriptional repression, modulation of signaling pathways, induction of hypertrophy, fibrosis, inflammation, oxidative stress, and interaction with other cardiogenic regulators [59, 60].

#### 8. MYOCD

##### Effect of TGF-beta on this Cardiogenic Regulator

MYOCD (Myocardin) is a transcriptional coactivator that plays a crucial role in cardiac development and function. It regulates the expression of genes involved in smooth and cardiac muscle differentiation, including those encoding contractile proteins and structural components necessary for cardiomyocyte integrity and function [61]. In adult cardiomyocytes, MYOCD continues to be involved in maintaining contractile function and responding to physiological and pathological stimuli. TGF-β impacts MYOCD in cardiomyocytes through various mechanisms. One key mechanism is transcriptional repression. TGF-β signaling activates Smad proteins, which can directly interact with the MYOCD promoter or regulatory regions, leading to reduced expression of MYOCD in cardiomyocytes. Additionally, TGF-β signaling can crosstalk with other pathways involved in cardiac remodeling and repair, such as BMP signaling, synergistically inhibiting MYOCD expression or activity through complex regulatory mechanisms. TGF-β is also a key inducer of fibrosis in the heart, promoting the differentiation of fibroblasts into myofibroblasts and increasing extracellular matrix (ECM) deposition. The resulting fibrotic environment alters cardiomyocyte interactions and signaling, indirectly impacting MYOCD function and regulatory networks. Moreover, TGF-β stimulates the production of pro-inflammatory cytokines and chemokines, contributing to chronic inflammation in the heart. This inflammatory milieu can disrupt MYOCD expression and function, impairing its ability to regulate cardiomyocyte gene expression essential for cardiac repair [62]. Furthermore, TGF-β signaling pathways include components that promote apoptosis in cardiomyocytes under conditions of stress or injury. Increased apoptosis can lead to reduced MYOCD expression and function, diminishing its role in maintaining cardiomyocyte integrity and function. TGF-β may also interact with other transcription factors or regulatory proteins involved in cardiomyocyte development and function, such as GATA4 or Mef2c. These interactions can form inhibitory complexes that suppress MYOCD activity or expression levels, further compromising its function in cardiac repair. TGF-β disrupts MYOCD function in cardiomyocytes during cardiac remodeling post-ischemic injury through multiple mechanisms, including transcriptional repression, modulation of signaling pathways, induction of fibrosis, inflammation, apoptosis, and interaction with other cardiogenic regulators [63, 64].

##### Effect of Angiotensin II on this Cardiogenic Regulator

MYOCD (Myocardin) is a critical transcriptional coactivator that regulates the expression of genes essential for cardiac muscle differentiation, contractility, and function. It plays a crucial role in cardiomyocyte development and contributes to the maintenance of cardiac structure and function in adults. Angiotensin II disrupts MYOCD function in cardiomyocytes through several mechanisms [65]. One key mechanism is the modulation of transcriptional activity.

Angiotensin II activates signaling pathways such as NF-κB and MAPK/ERK, which can lead to the recruitment of transcriptional repressors to the MYOCD gene promoter, decreasing MYOCD expression and disrupting its regulatory role in cardiomyocyte function and repair. Additionally, Angiotensin II interacts with the Renin-Angiotensin System (RAS), modulating intracellular signaling pathways that intersect with MYOCD-regulated pathways. This modulation can alter MYOCD transcriptional activity and its ability to regulate genes involved in cardiac adaptation and repair. Angiotensin II also induces hypertrophic signaling pathways (e.g., MAPK/ERK, PI3K/Akt) in cardiomyocytes, promoting hypertrophic growth. This altered cellular state may redirect resources away from regenerative processes regulated by MYOCD, hindering cardiac repair mechanisms [66]. Moreover, Angiotensin II promotes fibrosis by inducing the differentiation of fibroblasts into myofibroblasts, leading to increased extracellular matrix (ECM) deposition and fibrosis. The stiffened ECM can physically disrupt cellular interactions necessary for MYOCD function, impacting its regulatory role in cardiomyocyte physiology and repair. Angiotensin II also creates a pro-inflammatory environment by inducing cytokine production and immune cell activation. Chronic inflammation negatively impacts MYOCD expression and function, further impairing cardiac recovery post-ischemic injury. Furthermore, Angiotensin II induces oxidative stress in cardiomyocytes, leading to damage and functional impairment. This oxidative environment can alter the activity and stability of MYOCD, affecting its ability to regulate gene expression critical for cardiac adaptation and repair. Angiotensin II may also interact with other transcription factors or regulatory proteins involved in cardiomyocyte development and function, such as GATA4 or Mef2c. These interactions can form inhibitory complexes that suppress MYOCD activity or expression levels, further compromising its function in cardiac repair. Angiotensin II disrupts MYOCD function in cardiomyocytes during cardiac remodeling post-ischemic injury through multiple interconnected mechanisms, including transcriptional repression, modulation of signaling pathways, induction of hypertrophy, fibrosis, inflammation, oxidative stress, and interaction with other cardiogenic regulators [67, 68].

#### 9. MSX2

##### Effect of TGF-beta on this Cardiogenic Regulator

MSX2 is a transcription factor involved in cardiac development and tissue patterning. It regulates various genes essential for cell proliferation, differentiation, and ECM remodeling during embryonic heart development. In adult cardiomyocytes, MSX2 continues to be expressed and plays a role in cardiac remodeling and response to injury. TGF-β disrupts MSX2 function in cardiomyocytes through several mechanisms. One key mechanism is transcriptional repression, where TGF-β signaling activates Smad proteins, which can directly interact with the MSX2 gene promoter or regulatory regions. This interaction leads to transcriptional repression of MSX2, reducing its expression in cardiomyocytes [69]. Additionally, TGF-β signaling can crosstalk with other pathways involved in cardiac remodeling, such as BMP signaling, which interacts with MSX2. This interaction may synergistically inhibit MSX2 expression or activity through complex regulatory mechanisms. TGF-β also induces fibrosis in the heart, promoting the differentiation of fibroblasts into myofibroblasts and increasing ECM deposition. The fibrotic environment alters cardiomyocyte interactions and signaling, indirectly impacting MSX2 function and regulatory networks. Moreover, TGF-β stimulates the production of pro-inflammatory cytokines and chemokines, contributing to chronic inflammation in the heart [70]. This inflammatory milieu can disrupt MSX2 expression and function, impairing its ability to regulate cardiomyocyte gene expression critical for cardiac repair. TGF-β signaling pathways also include components that promote apoptosis in cardiomyocytes under conditions of stress or injury. Increased apoptosis can lead to reduced MSX2 expression and function, diminishing its role in maintaining cardiomyocyte integrity and function. Furthermore, TGF-β may interact with other transcription factors or regulatory proteins involved in cardiomyocyte development and function, such as GATA4 or Mef2c. These interactions can form inhibitory complexes that suppress MSX2 activity or expression levels, further compromising its function in cardiac repair. TGF-β disrupts MSX2 function in cardiomyocytes during cardiac remodeling post-ischemic injury through multiple mechanisms, including transcriptional repression, modulation of signaling pathways, induction of fibrosis, inflammation, apoptosis, and interaction with other cardiogenic regulators [71, 72].

##### Effect of Angiotensin II on this Cardiogenic Regulator

Angiotensin II disrupts MSX2 function in cardiomyocytes during cardiac remodeling post-ischemic injury through multiple interconnected mechanisms. One of the key mechanisms is transcriptional repression. Angiotensin II activates signaling pathways such as NF-κB and MAPK/ERK, which can lead to the recruitment of transcriptional repressors to the MSX2 gene promoter. This repression decreases MSX2 expression, disrupting its regulatory role in cardiomyocyte function and repair [73]. Additionally, Angiotensin II, a key component of the renin-angiotensin system (RAS), modulates intracellular signaling pathways that intersect with MSX2-regulated pathways. This modulation may alter MSX2 transcriptional activity and its ability to regulate genes involved in cardiac adaptation and repair.

Angiotensin II also induces a hypertrophic response by activating hypertrophic signaling pathways (e.g., MAPK/ERK, PI3K/Akt) in cardiomyocytes, promoting hypertrophic growth. This altered cellular state may redirect resources away from regenerative processes regulated by MSX2, thereby hindering cardiac repair mechanisms. Furthermore, Angiotensin II promotes fibrosis by inducing differentiation of fibroblasts into myofibroblasts, leading to increased extracellular matrix (ECM) deposition and fibrosis [74]. The stiffened ECM can physically disrupt cellular interactions necessary for MSX2 function, impacting its regulatory role in cardiomyocyte physiology and repair. The pro-inflammatory effects of Angiotensin II also contribute to the disruption of MSX2 function. Angiotensin II promotes a pro-inflammatory environment by inducing cytokine production and immune cell activation. Chronic inflammation negatively impacts MSX2 expression and function, further impairing cardiac recovery post-ischemic injury.

Additionally, Angiotensin II induces oxidative stress in cardiomyocytes, leading to damage and functional impairment. This oxidative environment can alter the activity and stability of MSX2, affecting its ability to regulate gene expression critical for cardiac adaptation and repair. Finally, Angiotensin II may interact with other transcription factors or regulatory proteins involved in cardiomyocyte development and function, such as GATA4 or Mef2c. These interactions can form inhibitory complexes that suppress MSX2 activity or expression levels, further compromising its function in cardiac repair [75, 76].

#### 10. HOPX

##### Effect of TGF-beta on this Cardiogenic Regulator

TGF-β disrupts HOPX function in cardiomyocytes during cardiac remodeling post-ischemic injury through multiple mechanisms. HOPX (Homeodomain-only protein X) is a transcriptional regulator involved in cardiac development, maintenance of cardiomyocyte quiescence, and response to stress. It plays a critical role in regulating gene expression networks that influence cardiomyocyte proliferation, differentiation, and function [77]. TGF-β signaling activates Smad proteins, which can directly bind to the HOPX promoter and inhibit its transcription, leading to reduced HOPX expression in cardiomyocytes and disrupting its regulatory role in maintaining quiescence and function. Additionally, TGF-β signaling can crosstalk with other pathways involved in cardiac remodeling and repair, potentially altering HOPX transcriptional activity or stability and influencing its ability to regulate genes critical for cardiomyocyte homeostasis and repair [78]. TGF-β also promotes fibrosis in the heart by stimulating the differentiation of fibroblasts into myofibroblasts and increasing extracellular matrix (ECM) deposition. The fibrotic environment can disrupt cardiomyocyte interactions and signaling, indirectly impacting HOPX function and regulatory networks. Furthermore, TGF-β induces a pro-inflammatory milieu by stimulating the production of cytokines and chemokines, and chronic inflammation negatively affects HOPX expression and function in cardiomyocytes, potentially compromising its role in maintaining cardiac homeostasis. TGF-β signaling pathways include components that promote apoptosis in cardiomyocytes under stress conditions, and increased apoptosis can lead to reduced HOPX expression, impairing its ability to regulate cardiomyocyte survival and function [79]. TGF-β may interact with other transcription factors or regulatory proteins involved in cardiomyocyte development and function, such as GATA4 or Mef2c. These interactions can form inhibitory complexes that suppress HOPX activity or expression levels, further compromising its function in cardiac repair. Understanding these mechanisms provides insights into how TGF-β contributes to impaired cardiac repair and regeneration, suggesting potential therapeutic targets to enhance HOPX-mediated recovery in ischemic heart disease [80].

##### Effect of Angiotensin II on this Cardiogenic Regulator

Investigating how Angiotensin II impacts HOPX in cardiomyocytes during cardiac remodeling post-ischemic injury involves considering several potential mechanisms. Angiotensin II activates signaling pathways such as NF-κB and MAPK/ERK, leading to the recruitment of transcriptional repressors to the HOPX gene promoter. This repression decreases HOPX expression, disrupting its regulatory role in maintaining cardiomyocyte quiescence and function [81].

As a key component of the Renin-Angiotensin System (RAS), Angiotensin II modulates intracellular signaling pathways that intersect with HOPX-regulated pathways, potentially altering HOPX transcriptional activity and its ability to regulate genes involved in cardiac adaptation and repair. Angiotensin II activates hypertrophic signaling pathways (e.g., MAPK/ERK, PI3K/Akt) in cardiomyocytes, promoting hypertrophic growth. This altered cellular state may divert resources away from regulatory processes governed by HOPX, thereby hindering its role in maintaining cardiomyocyte quiescence and function. Additionally, Angiotensin II induces differentiation of fibroblasts into myofibroblasts, leading to increased extracellular matrix (ECM) deposition and fibrosis [82]. The altered ECM can disrupt cardiomyocyte interactions and signaling necessary for HOPX function, impacting its regulatory role in cardiomyocyte physiology and repair. Angiotensin II promotes a pro-inflammatory environment by inducing cytokine production and immune cell activation, and chronic inflammation negatively impacts HOPX expression and function in cardiomyocytes, potentially compromising its ability to maintain cardiac homeostasis. Furthermore, Angiotensin II induces oxidative stress in cardiomyocytes, leading to damage and functional impairment. This oxidative environment can alter the activity and stability of HOPX, affecting its ability to regulate gene expression critical for cardiac adaptation and repair. Angiotensin II may also interact with other transcription factors or regulatory proteins involved in cardiomyocyte development and function, such as GATA4 or Mef2c. These interactions can form inhibitory complexes that suppress HOPX activity or expression levels, further compromising its function in cardiac repair.

Angiotensin II disrupts HOPX function in cardiomyocytes during cardiac remodeling post-ischemic injury through multiple interconnected mechanisms, including transcriptional repression, modulation of signaling pathways, induction of hypertrophy, fibrosis, inflammation, oxidative stress, and interaction with other cardiogenic regulators [83, 84].

#### 11. Wnt-signaling pathway

##### Effect of TGF-beta on this Cardiogenic Regulator

Wnt signaling is crucial for various aspects of cardiac development, including cardiomyocyte proliferation, differentiation, and morphogenesis. It regulates gene expression programs that are essential for maintaining cardiac structure and function. TGF-β signaling can inhibit the expression of Wnt ligands such as Wnt1, Wnt3a, and Wnt5a, which are critical for activating canonical and non-canonical Wnt pathways in cardiomyocytes [85]. This downregulation limits the activation of Wnt signaling pathways necessary for cardiomyocyte proliferation and differentiation during cardiac repair. TGF-β can influence the expression levels or activity of Frizzled receptors, which are necessary for Wnt ligand binding and signal transduction. Changes in Frizzled receptor expression or function can impair Wnt signaling activation in cardiomyocytes, impacting their ability to undergo repair processes post-ischemic injury. TGF-β signaling can promote the degradation of β-catenin, a key mediator of canonical Wnt signaling. Reduced β-catenin levels in cardiomyocytes hinder the activation of Wnt target genes involved in cell survival, proliferation, and tissue repair. TGF-β induces the production of extracellular matrix (ECM) proteins and promotes fibroblast activation, leading to ECM remodeling and fibrosis [86]. The altered ECM composition can physically disrupt Wnt signaling activation and transmission in cardiomyocytes, impairing their ability to respond to repair signals. TGF-β signaling can cross-talk with BMP (Bone Morphogenetic Protein) signaling pathways, which interact with Wnt pathways during cardiac development and repair. Dysregulation of BMP signaling by TGF-β can indirectly affect Wnt pathway activity in cardiomyocytes, contributing to impaired repair mechanisms. TGF-β promotes a pro-inflammatory environment in the heart by inducing cytokine production and immune cell activation. Chronic inflammation can disrupt Wnt signaling activation and maintenance in cardiomyocytes, limiting their ability to initiate repair processes post-ischemic injury. TGF-β disrupts Wnt signaling in cardiomyocytes during cardiac remodeling post-ischemic injury through multiple mechanisms, including transcriptional downregulation of Wnt ligands, modulation of Wnt co-receptors, inhibition of β-catenin pathway, induction of fibrosis, cross-talk with other signaling pathways, and modulation of inflammatory responses [87, 88].

##### Effect of Angiotensin II on this Cardiogenic Regulator

Angiotensin II can influence the expression and secretion of Wnt ligands such as Wnt1, Wnt3a, and Wnt5a. Reduced levels of these ligands impair the activation of Wnt signaling pathways critical for cardiomyocyte proliferation and differentiation during cardiac repair. Angiotensin II may also affect the expression levels or activity of Frizzled receptors, which are necessary for Wnt ligand binding and signal transduction [89]. Changes in Frizzled receptor expression or function can impair Wnt signaling activation in cardiomyocytes, impacting their ability to undergo repair processes post-ischemic injury. Angiotensin II signaling pathways can promote the degradation of β-catenin, a key mediator of canonical Wnt signaling. Reduced β-catenin levels in cardiomyocytes hinder the activation of Wnt target genes involved in cell survival, proliferation, and tissue repair. Angiotensin II activates hypertrophic signaling pathways (e.g., MAPK/ERK, PI3K/Akt) in cardiomyocytes, promoting hypertrophic growth. This altered cellular state may divert resources away from regulatory processes governed by Wnt signaling, thereby hindering its role in maintaining cardiomyocyte function and repair [90]. Angiotensin II induces differentiation of fibroblasts into myofibroblasts, leading to increased extracellular matrix (ECM) deposition and fibrosis. The altered ECM composition can physically disrupt Wnt signaling activation and transmission in cardiomyocytes, impairing their ability to respond to repair signals. Angiotensin II promotes a pro-inflammatory environment in the heart by inducing cytokine production and immune cell activation [91]. Chronic inflammation can disrupt Wnt signaling activation and maintenance in cardiomyocytes, limiting their ability to initiate repair processes post-ischemic injury. Angiotensin II also induces oxidative stress in cardiomyocytes, leading to damage and functional impairment. This oxidative environment can further impair Wnt signaling activity, affecting its role in cardiomyocyte survival and repair mechanisms. Angiotensin II disrupts Wnt signaling in cardiomyocytes during cardiac remodeling post-ischemic injury through multiple mechanisms, including modulation of Wnt ligands and receptors, inhibition of the β-catenin pathway, induction of hypertrophy and fibrosis, and interaction with inflammatory and oxidative stress pathways [92].

#### 12. Notch Signaling Pathway

##### Effect of TGF-beta on this Cardiogenic Regulator

Notch signaling is crucial for various aspects of cardiac development, including cardiomyocyte differentiation, proliferation, and maturation. It regulates cell fate decisions and maintains cardiac homeostasis by influencing gene expression programs. TGF-β signaling can inhibit the expression of Notch receptors (Notch1-4) and ligands (Delta-like [DLL] and Jagged [JAG] families) in cardiomyocytes [93]. Reduced levels of these receptors and ligands impair Notch signaling activation, disrupting its regulatory role in cardiomyocyte differentiation and function. TGF-β can influence the stability and activity of NICD, the active form of Notch signaling, by promoting its degradation or inhibiting its cleavage. This modulation limits NICD-mediated transcriptional activation of downstream target genes critical for cardiac repair processes. TGF-β promotes cardiac fibrosis by stimulating ECM production and fibroblast activation. The fibrotic environment alters the cellular interactions necessary for Notch signaling activation and propagation in cardiomyocytes, impairing its role in regulating cell fate and function. TGF-β signaling can cross-talk with BMP pathways, which regulate Notch signaling during cardiac development and repair. Dysregulation of BMP signaling by TGF-β can indirectly affect Notch pathway activity in cardiomyocytes, contributing to impaired repair mechanisms. TGF-β promotes a pro-inflammatory milieu in the heart by inducing cytokine production and immune cell activation [94]. Chronic inflammation disrupts Notch signaling activation and maintenance in cardiomyocytes, limiting their ability to initiate repair processes post-ischemic injury. TGF-β disrupts Notch signaling in cardiomyocytes during cardiac remodeling post-ischemic injury through multiple mechanisms, including transcriptional downregulation of Notch receptors and ligands, modulation of Notch pathway components like NICD stability, induction of fibrosis, cross-talk with other signaling pathways, and modulation of inflammatory responses [95, 96].

##### Effect of Angiotensin II on this Cardiogenic Regulator

Angiotensin II can influence the expression and activation of Notch receptors (Notch1-4) in cardiomyocytes. Reduced levels of these receptors impair Notch signaling activation, affecting its regulatory role in cardiomyocyte differentiation, proliferation, and function during cardiac repair. Angiotensin II may also affect the expression or activity of Notch ligands (Delta-like [DLL] and Jagged [JAG] families) necessary for ligand-receptor interactions.

Changes in ligand availability can disrupt Notch signaling pathway activation and downstream effects on cardiomyocyte behavior and repair processes. Angiotensin II signaling pathways can promote the degradation or inhibit the cleavage of Notch Intracellular Domain (NICD), the active form of Notch signaling. Reduced NICD stability limits its transcriptional activation of downstream target genes critical for cardiac repair and regeneration [97].

Angiotensin II activates hypertrophic signaling pathways (e.g., MAPK/ERK, PI3K/Akt) in cardiomyocytes, promoting hypertrophic growth and altering cellular functions. This hypertrophic state may interfere with Notch signaling-mediated regulation of cardiomyocyte growth and repair mechanisms. Additionally, Angiotensin II induces cardiac fibrosis by stimulating ECM production and fibroblast activation [98]. The fibrotic environment disrupts cellular interactions necessary for Notch signaling activation and propagation in cardiomyocytes, impairing its role in maintaining cardiac homeostasis and repair. Angiotensin II promotes a pro-inflammatory environment in the heart by inducing cytokine production and immune cell activation. Chronic inflammation disrupts Notch signaling activation and maintenance in cardiomyocytes, limiting their ability to initiate repair processes post-ischemic injury.

Furthermore, Angiotensin II induces oxidative stress in cardiomyocytes, leading to cellular damage and functional impairment. This oxidative environment can further impair Notch signaling activity, affecting its role in cardiomyocyte survival and repair mechanisms. Angiotensin II disrupts Notch signaling in cardiomyocytes during cardiac remodeling post-ischemic injury through multiple mechanisms, including modulation of Notch receptors and ligands, inhibition of NICD stability, induction of hypertrophic response and fibrosis, and interaction with inflammatory and oxidative stress pathways [99, 100].

#### 13. FGF Signaling Pathway

##### Effect of TGF-beta on this Cardiogenic Regulator

TGF-beta exerts significant influence on FGF signaling in cardiomyocytes, impacting various aspects of cardiac remodeling post-ischemic injury through several interconnected mechanisms. TGF-beta signaling can alter the expression and activation of FGF receptors (FGFRs) in cardiomyocytes, leading to reduced responsiveness to FGF ligands such as FGF1, FGF2, and FGF23. This downregulation impairs FGF signaling pathways critical for cardiomyocyte proliferation, survival, and repair [101]. Additionally, TGF-beta may directly affect the expression or secretion of FGF ligands, disrupting the activation and propagation of FGF signaling in cardiomyocytes and thereby compromising their function and repair processes. Furthermore, TGF-beta interferes with intracellular signaling pathways essential for FGF-mediated effects. For instance, FGF signaling typically activates the MAPK/ERK pathway, crucial for cardiomyocyte survival and proliferation. TGF-beta signaling can inhibit or cross-regulate MAPK/ERK activation, diminishing FGF-mediated cardioprotective effects during cardiac repair. TGF-beta also plays a significant role in promoting cardiac fibrosis by stimulating extracellular matrix (ECM) production and activating fibroblasts. This fibrotic environment disrupts the cellular interactions necessary for proper FGF signaling activation and transmission in cardiomyocytes, thereby impairing its role in promoting cardiomyocyte survival and repair mechanisms. Moreover, TGF-beta signaling interacts with other pathways such as BMP (Bone Morphogenetic Protein), which also influence FGF signaling in cardiac development and repair [102]. Dysregulation of BMP signaling by TGF-beta indirectly affects FGF pathway activity in cardiomyocytes, contributing further to impaired repair mechanisms post-ischemic injury. Additionally, TGF-beta promotes a pro-inflammatory environment in the heart by inducing cytokine production and immune cell activation. Chronic inflammation disrupts the activation and maintenance of FGF signaling in cardiomyocytes, limiting their ability to initiate and sustain repair processes essential for recovering from ischemic injury. TGF-beta disrupts FGF signaling in cardiomyocytes during cardiac remodeling post-ischemic injury through multiple mechanisms, including modulation of FGF receptors and ligands, interference with intracellular signaling pathways like MAPK/ERK, promotion of fibrosis, cross-talk with other signaling pathways such as BMP, and modulation of inflammatory responses. These findings highlight the complex interactions between TGF-beta and FGF signaling pathways in cardiac pathology, suggesting potential therapeutic targets to enhance cardiomyocyte repair and recovery following ischemic events [103, 104].

##### Effect of Angiotensin II on this Cardiogenic Regulator

Angiotensin II exerts significant effects on FGF signaling in cardiomyocytes during cardiac remodeling post-ischemic injury through a variety of interconnected mechanisms. Angiotensin II influences the expression and activation of FGF receptors (FGFRs) in cardiomyocytes, leading to reduced responsiveness to FGF ligands such as FGF1, FGF2, and FGF23. This downregulation impairs FGF signaling pathways critical for cardiomyocyte survival, proliferation, and repair [105]. Additionally, Angiotensin II can alter the expression or secretion of FGF ligands, disrupting the activation and propagation of FGF signaling in cardiomyocytes and compromising their function and repair processes. Moreover, Angiotensin II interferes with intracellular signaling pathways crucial for FGF-mediated effects. For instance, FGF signaling typically activates the MAPK/ERK pathway, essential for cardiomyocyte survival and proliferation.

Angiotensin II signaling may inhibit or cross-regulate MAPK/ERK activation, thereby diminishing FGF-mediated cardioprotective effects during cardiac repair. Angiotensin II also induces hypertrophic signaling pathways like MAPK/ERK and PI3K/Akt in cardiomyocytes, promoting hypertrophic growth and altering cellular functions [106]. This hypertrophic state may interfere with FGF signaling-mediated regulation of cardiomyocyte growth and repair mechanisms. Additionally, Angiotensin II promotes cardiac fibrosis by stimulating extracellular matrix (ECM) production and activating fibroblasts. The resulting fibrotic environment disrupts cellular interactions necessary for proper FGF signaling activation and transmission in cardiomyocytes, impairing its role in promoting cardiomyocyte survival and repair mechanisms. Furthermore, Angiotensin II signaling interacts with other pathways such as TGF-β, which also play significant roles in cardiac remodeling and fibrosis. Dysregulation of TGF-β signaling by Angiotensin II indirectly affects FGF pathway activity in cardiomyocytes, further contributing to impaired repair mechanisms post-ischemic injury. Additionally, Angiotensin II induces oxidative stress in cardiomyocytes, leading to cellular damage and functional impairment. This oxidative environment further impairs FGF signaling activity, affecting its crucial role in cardiomyocyte survival and repair mechanisms [107]. Moreover, Angiotensin II promotes a pro-inflammatory environment in the heart by inducing cytokine production and immune cell activation. Chronic inflammation disrupts FGF signaling activation and maintenance in cardiomyocytes, limiting their ability to initiate and sustain repair processes essential for recovering from ischemic injury. Angiotensin II disrupts FGF signaling in cardiomyocytes during cardiac remodeling post-ischemic injury through multiple complex mechanisms, including modulation of FGF receptors and ligands, interference with intracellular signaling pathways like MAPK/ERK, promotion of hypertrophic and fibrotic responses, cross-talk with other signaling pathways like TGF-β, and modulation of oxidative stress and inflammatory responses. These findings point to the complex interplay between Angiotensin II and FGF signaling pathways in cardiac pathophysiology, suggesting potential therapeutic targets to enhance cardiomyocyte repair and recovery following ischemic events [108].

#### 14. BMPs

##### Effect of TGF-beta on this Cardiogenic Regulator

TGF-beta significantly impacts BMP signaling in cardiomyocytes during cardiac remodeling post-ischemic injury through several interconnected mechanisms. TGF-beta influences the expression and activation of BMP receptors such as BMPR1A, BMPR1B, and BMPR2 in cardiomyocytes. This modulation reduces the responsiveness of cardiomyocytes to BMP ligands like BMP2, BMP4, and BMP7, disrupting downstream signaling pathways crucial for cardiomyocyte differentiation, proliferation, and repair processes. Additionally, TGF-beta can alter the expression or secretion of BMP ligands within cardiomyocytes, further impairing BMP signaling pathway activation and its beneficial effects on cardiomyocyte function and repair [109]. Moreover, TGF-beta interferes with intracellular signaling pathways essential for BMP-mediated effects. BMP signaling typically activates Smad-dependent pathways such as Smad1/5/8, which regulate gene expression involved in cardiomyocyte differentiation and survival. TGF-beta signaling may interfere with Smad activation or promote cross-inhibition, thereby diminishing BMP’s ability to promote cardiomyocyte repair during cardiac remodeling. TGF-beta also plays a critical role in promoting cardiac fibrosis by stimulating extracellular matrix (ECM) production and activating fibroblasts. The resulting fibrotic environment disrupts the cellular interactions necessary for proper BMP signaling activation and transmission in cardiomyocytes, impairing its role in promoting cardiomyocyte survival and repair mechanisms post-ischemic injury. Furthermore, TGF-beta signaling can cross-talk with other signaling pathways such as the Wnt pathway, which also regulates BMP signaling during cardiac development and repair [110]. Dysregulation of Wnt signaling by TGF-beta indirectly affects BMP pathway activity in cardiomyocytes, contributing further to impaired repair mechanisms. Additionally, TGF-beta promotes a pro-inflammatory environment in the heart by inducing cytokine production and immune cell activation. Chronic inflammation disrupts BMP signaling activation and maintenance in cardiomyocytes, limiting their ability to initiate and sustain repair processes essential for recovering from ischemic injury. TGF-beta disrupts BMP signaling in cardiomyocytes during cardiac remodeling post-ischemic injury through multiple complex mechanisms, including modulation of BMP receptors and ligands, interference with intracellular Smad signaling pathways, promotion of fibrosis, cross-talk with other signaling pathways like Wnt, and modulation of inflammatory responses [111, 112].

##### Effect of Angiotensin II on this Cardiogenic Regulator

Angiotensin II exerts a significant impact on BMP signaling in cardiomyocytes during cardiac remodeling post-ischemic injury through various interconnected mechanisms. It influences the expression and activation of BMP receptors such as BMPR1A, BMPR1B, and BMPR2 in cardiomyocytes. By reducing the levels of these receptors, Angiotensin II impairs the responsiveness of cardiomyocytes to BMP ligands like BMP2, BMP4, and BMP7, disrupting downstream signaling pathways critical for cardiomyocyte differentiation, proliferation, and repair processes [113]. Additionally, Angiotensin II can alter the expression or secretion of BMP ligands within cardiomyocytes, further undermining BMP signaling pathway activation and its beneficial effects on cardiomyocyte function and repair. Moreover, Angiotensin II interferes with intracellular signaling pathways essential for BMP-mediated effects. BMP signaling typically activates Smad-dependent pathways such as Smad1/5/8, which regulate gene expression involved in cardiomyocyte differentiation and survival. Angiotensin II signaling may interfere with Smad activation or promote cross-inhibition, thereby diminishing BMP’s ability to promote effective cardiomyocyte repair during cardiac remodeling [114]. Furthermore, Angiotensin II induces hypertrophic signaling pathways in cardiomyocytes, such as MAPK/ERK and PI3K/Akt, which promote hypertrophic growth and alter cellular functions. This hypertrophic state can interfere with BMP signaling-mediated regulation of cardiomyocyte growth and repair mechanisms. Additionally, Angiotensin II promotes cardiac fibrosis by stimulating extracellular matrix (ECM) production and activating fibroblasts. The resulting fibrotic environment disrupts the cellular interactions necessary for proper BMP signaling activation and propagation in cardiomyocytes, impairing its role in promoting cardiomyocyte survival and repair mechanisms post-ischemic injury. Angiotensin II also interacts with other signaling pathways like the TGF-beta pathway, which also plays critical roles in cardiac remodeling and fibrosis. Dysregulation of TGF-beta signaling by Angiotensin II indirectly affects BMP pathway activity in cardiomyocytes, contributing further to impaired repair mechanisms [115]. Additionally, Angiotensin II induces oxidative stress in cardiomyocytes, leading to cellular damage and functional impairment. This oxidative environment further impairs BMP signaling activity, thereby affecting its crucial role in cardiomyocyte survival and repair mechanisms. Furthermore, Angiotensin II promotes a pro-inflammatory environment in the heart by inducing cytokine production and immune cell activation. Chronic inflammation disrupts BMP signaling activation and maintenance in cardiomyocytes, limiting their ability to initiate and sustain repair processes essential for recovering from ischemic injury. Angiotensin II disrupts BMP signaling in cardiomyocytes during cardiac remodeling post-ischemic injury through multiple complex mechanisms. These include modulation of BMP receptors and ligands, interference with intracellular Smad signaling pathways, promotion of hypertrophic and fibrotic responses, cross-talk with other signaling pathways like TGF-beta, and modulation of oxidative stress and inflammatory responses. Understanding these interactions could inform potential therapeutic strategies aimed at preserving BMP signaling integrity and enhancing cardiac repair mechanisms in ischemic heart disease [116, 117].

## Discussion

### Cardiac Remodeling Post-Ischemic Injury: Disruption of Cardiac Proliferation vs Differentiation

#### 1. Isl1 (Islet1)

Role: Isl1 is critical for progenitor cell proliferation and differentiation during cardiac development.

Impact of Ischemic Injury: Ischemic injury may reduce Isl1 expression or alter its regulatory networks, impairing progenitor cell activation and differentiation in damaged myocardium.

Consequence: This disruption can hinder the recruitment and differentiation of progenitor cells needed for myocardial repair and regeneration post-ischemic injury.

#### 2. Brg1/Baf60 – Smarcd3 Complex

Role: Involved in chromatin remodeling and gene expression regulation, influencing cardiac differentiation pathways. Impact of Ischemic Injury: Ischemic damage can dysregulate Brg1/Baf60 complex activity, leading to altered gene expression profiles crucial for cardiomyocyte differentiation and maturation.

Consequence: Impaired chromatin remodeling may hinder the transition of progenitor cells or immature cardiomyocytes into functional, differentiated cardiomyocytes, limiting repair capacity.

#### 3. Nkx2–5

Role: Essential transcription factor for cardiomyocyte differentiation and maturation.

Impact of Ischemic Injury: Ischemic conditions may disrupt Nkx2–5 expression or function, affecting the proper development and maturation of cardiomyocytes in damaged areas.

Consequence: Reduced Nkx2–5 activity can impair the differentiation of progenitor cells into mature cardiomyocytes, contributing to ineffective myocardial repair and remodeling.

#### 4. GATA4

Role: Transcription factor involved in early cardiac development and maintenance of cardiomyocyte function. Impact of Ischemic Injury: Ischemic injury may alter GATA4 expression levels or its regulatory pathways, compromising cardiomyocyte survival and function.

Consequence: Disrupted GATA4 function can impair both proliferation and differentiation processes in cardiomyocytes, affecting their ability to respond to injury and contribute to tissue repair.

#### 5. Tbx5

Role: Essential for heart development and regulation of chamber-specific gene expression.

Impact of Ischemic Injury: Ischemic conditions can disrupt Tbx5 expression or activity, affecting chamber specification and differentiation processes.

Consequence: Altered Tbx5 function may lead to improper differentiation of cardiomyocytes or progenitor cells, impairing myocardial remodeling and repair post-ischemic injury.

#### 6. Mef2c

Role: Transcription factor critical for cardiomyocyte differentiation, growth, and survival.

Impact of Ischemic Injury: Ischemic damage may perturb Mef2c signaling pathways, affecting its role in regulating cardiomyocyte differentiation and survival.

Consequence: Dysregulated Mef2c activity can impair the differentiation of progenitor cells into functional cardiomyocytes, diminishing the heart’s capacity for repair and regeneration.

#### 7. HAND1/2

Role: Transcription factors involved in heart development and differentiation of specific cardiac cell types.

Impact of Ischemic Injury: Ischemic injury may disrupt HAND1/2 expression or function, impairing the differentiation of progenitor cells into specialized cardiac cell lineages.

Consequence: Altered HAND1/2 activity can affect the diversity and function of cardiac cells formed during repair processes, potentially limiting effective tissue remodeling post-ischemic injury.

#### 8. MYOCD (Myocardin)

Role: Co-activator of serum response factor (SRF) involved in cardiac muscle gene expression and differentiation. Impact of Ischemic Injury: Ischemic conditions can disturb MYOCD expression or its interaction with SRF, affecting cardiomyocyte differentiation and contractile function.

Consequence: Impaired MYOCD-SRF signaling can hinder the maturation of cardiomyocytes from progenitor cells, reducing the functional capacity of regenerated myocardium.

#### 9. MSX2

Role: Regulator of cell fate decisions and differentiation during heart development.

Impact of Ischemic Injury: Ischemic injury may alter MSX2 expression or activity, influencing cell fate determination and differentiation pathways.

Consequence: Disrupted MSX2 function can lead to aberrant differentiation of progenitor cells or immature cardiomyocytes, affecting the structural and functional integrity of regenerated myocardium.

#### 10. HOPX

Role: Regulator of cardiac progenitor cell proliferation and differentiation.

Impact of Ischemic Injury: Ischemic damage can perturb HOPX expression or function, impairing its role in regulating progenitor cell behaviors during cardiac repair.

Consequence: Altered HOPX activity may lead to dysregulated progenitor cell proliferation or differentiation, limiting effective myocardial regeneration post-ischemic injury.

#### 11. Wnt-Signaling Pathway

Role: Critical for cardiac development, proliferation, and differentiation.

Impact of Ischemic Injury: Ischemic conditions can dysregulate Wnt signaling pathways, affecting progenitor cell fate decisions and cardiomyocyte differentiation.

Consequence: Impaired Wnt signaling may hinder the proper differentiation of progenitor cells into functional cardiomyocytes, compromising myocardial repair and regeneration.

#### 12. Notch Signaling

Role: Involved in cell fate determination and differentiation during cardiac development.

Impact of Ischemic Injury: Ischemic injury may disrupt Notch signaling pathways, affecting progenitor cell fate decisions and cardiomyocyte differentiation.

Consequence: Altered Notch signaling can lead to aberrant differentiation patterns or impaired cell-cell communication, compromising effective myocardial repair post-ischemic injury.

#### 13. FGF (Fibroblast Growth Factor)

Role: Involved in cardiac growth, angiogenesis, and cardiomyocyte survival.

Impact of Ischemic Injury: Ischemic conditions may alter FGF signaling, affecting angiogenic responses and cardiomyocyte survival pathways.

Consequence: Dysregulated FGF signaling can impair angiogenesis and cardiomyocyte survival, limiting effective tissue repair and regeneration post-ischemic injury.

#### 14. BMPs (Bone Morphogenetic Proteins)

Role: Critical for cardiac development, differentiation, and repair.

Impact of Ischemic Injury: Ischemic damage can disrupt BMP signaling pathways, affecting progenitor cell differentiation and cardiomyocyte maturation.

Consequence: Impaired BMP signaling may hinder the differentiation of progenitor cells into functional cardiomyocytes, compromising myocardial repair and regeneration processes.

Cardiac remodeling post-ischemic injury disrupts the delicate balance between proliferation and differentiation roles of cardiogenic regulators. Understanding these disruptions is crucial for developing targeted therapies aimed at enhancing cardiac repair and regeneration, thereby improving clinical outcomes for patients with ischemic heart disease.

#### TGF-beta and Angiotensin II as key regulators of cardiac remodeling

While numerous regulators influence cardiac remodeling, TGF-β and Angiotensin II stand out due to their crucial roles in this complex process. TGF-β plays a crucial role by mediating fibrosis, affecting fibroblast activation, and influencing the deposition of extracellular matrix (ECM). It also modulates inflammatory responses and cellular differentiation, contributing to both reparative and pathological remodeling following ischemic injury. The signaling pathways of TGF-β are instrumental in the development of cardiac hypertrophy and fibrosis, further highlighting its significance in remodeling processes. Angiotensin II, a key component of the renin-angiotensin-aldosterone system (RAAS), exerts profound effects on cardiac cells. It induces cardiac hypertrophy, fibrosis, and inflammation through actions on cardiomyocytes, fibroblasts, and vascular cells. Moreover, Angiotensin II stimulates the production of pro-fibrotic and pro-inflammatory factors, including TGF-β, thereby creating a positive feedback loop that exacerbates cardiac remodeling. Additionally, it promotes oxidative stress and apoptosis, contributing to adverse structural and functional changes in the heart post-ischemic injury. The interactions between TGF-β and Angiotensin II further amplify their effects on cardiac remodeling. Angiotensin II can upregulate TGF-β expression, enhancing its fibrotic and hypertrophic effects. Both factors activate critical signaling pathways such as the Smad pathway (associated with TGF-β) and the MAPK pathway (linked to Angiotensin II), which converge to promote ECM deposition, differentiation of myofibroblasts, and hypertrophy of cardiomyocytes. This synergistic interplay points to the importance of targeting both TGF-β and Angiotensin II pathways for therapeutic intervention aimed at mitigating adverse cardiac remodeling and improving outcomes following ischemic injury. Given their central roles and the critical nature of their interactions, interventions that selectively target TGF-β and Angiotensin II pathways hold significant promise in clinical settings. These approaches have the potential to attenuate detrimental remodeling processes, thereby enhancing cardiac function and overall patient outcomes post-ischemic injury.

#### Significance of the study

Specific cardiogenic regulators such as Isl1, Nkx2–5, and GATA4 have been identified as targets disrupted by these molecules, elucidating their critical roles in myocardial repair and regeneration processes. The study highlights the crosstalk between TGF-beta and Angiotensin II signaling pathways. This interplay influences downstream pathways essential for cardiac remodeling, including Wnt-signaling, Notch, and BMPs, which collectively regulate cellular responses and tissue repair mechanisms in the heart. Such pathway interactions further emphasize the multidimensional nature of cardiac remodeling processes. Clinically, insights into these dysregulated mechanisms offer potential biomarkers for assessing disease severity in ischemic heart disease. These biomarkers could also guide personalized treatment strategies, moving beyond conventional approaches towards more targeted interventions tailored to individual patient needs. Furthermore, by delineating how TGF-beta and Angiotensin II disrupt cardiogenic regulators, the research identifies new therapeutic targets. These targets hold promise for mitigating adverse remodeling processes and improving cardiac function post-ischemic injury. This approach represents a significant shift towards developing precise molecular interventions that address the root causes of pathological cardiac remodeling. Overall, this research fills critical gaps in understanding the molecular underpinnings of pathological cardiac remodeling. It sets the stage for new therapeutic approaches aimed at enhancing outcomes for patients with ischemic heart disease, ultimately paving the way for improved patient care and quality of life.

This research addresses significant gaps in understanding the role of molecular disruptions, particularly focusing on TGF-beta and Angiotensin II, in contributing to pathological cardiac remodeling post-ischemic injury. Previous studies have broadly implicated TGF-beta and Angiotensin II in cardiac remodeling, emphasizing their involvement in processes such as fibrosis, hypertrophic responses, and inflammatory pathways. However, specific molecular mechanisms by which these factors disrupt cardiogenic regulators like Isl1, Nkx2–5, and GATA4 have remained inadequately understood until now. This research looks deeper into the regulatory networks affected by TGF-beta and Angiotensin II. It uncovers how these molecules influence critical aspects such as transcription factors, signaling pathways (e.g., Wnt-signaling, Notch, BMPs), and chromatin remodeling complexes (e.g., Brg1/Baf60 – Smarcd3 complex). By elucidating these disruptions, the study provides a more comprehensive view of how molecular changes drive pathological remodeling processes in the heart. Understanding these molecular disruptions is crucial for advancing beyond symptomatic treatments towards targeted molecular interventions. This approach facilitates the development of therapies aimed at modulating specific pathways or restoring normal regulatory functions affected by ischemic injury. Moreover, it opens avenues for identifying biomarkers that could predict disease progression or response to treatment, thereby enhancing personalized medicine approaches in cardiovascular care. By identifying key nodes in disrupted molecular pathways, such as potential drug targets or candidates for gene therapy, this research may contribute to new therapeutic strategies. These strategies may involve small molecule inhibitors, biologics targeting specific receptors or signaling molecules, or gene editing techniques to restore or enhance the function of cardiogenic regulators. The translational impact of these findings is substantial, not only deepening our fundamental understanding of cardiac remodeling but also directly influencing clinical practice. They provide a solid foundation for conducting clinical trials aimed at validating the efficacy and safety of targeted interventions designed to mitigate adverse remodeling processes in patients with ischemic heart disease. This research significantly advances our knowledge by elucidating how molecular disruptions contribute to pathological cardiac remodeling. It paves the way for new approaches to the treatment and management of ischemic heart disease, offering promise beyond traditional symptomatic relief towards more effective and tailored therapeutic interventions.

#### Clinical Implications

Insights into the dysregulated mechanisms of cardiac remodeling post-ischemic injury, particularly focusing on the roles of TGF-beta and Angiotensin II, have profound clinical implications.

1. **Biomarker Discovery**: The study identifies specific biomarkers associated with disrupted cardiogenic regulators such as Isl1, Nkx2–5, GATA4, and others. These biomarkers can serve as indicators of disease severity, progression, and response to treatment in patients with ischemic heart disease. For example, elevated levels of certain transcription factors or signaling molecules in blood or cardiac tissue samples may correlate with increased myocardial damage or poorer prognosis.
2. **Risk Stratification**: Biomarkers identified through this research can aid in risk stratification of patients following ischemic events. Clinicians can use these markers to assess individual risk profiles and tailor management strategies accordingly. For instance, patients identified with high-risk biomarker profiles may benefit from more aggressive monitoring or earlier intervention to prevent adverse remodeling and progression to heart failure.
3. **Personalized Treatment Approaches**: Understanding how TGF-beta and Angiotensin II disrupt cardiogenic regulators allows for the development of personalized treatment strategies. Therapies targeting specific molecular pathways or restoring normal regulatory functions could be tailored based on the patient’s biomarker profile. This approach enhances the efficacy of treatment while minimizing adverse effects by directly addressing the underlying molecular mechanisms driving cardiac remodeling.
4. **Monitoring Disease Progression**: Biomarkers can also serve as tools for monitoring disease progression over time. Serial measurements of biomarker levels can provide insights into the effectiveness of interventions and help adjust treatment plans as needed. This continuous monitoring improves patient outcomes by enabling timely adjustments to therapy in response to changes in disease activity or severity.
5. **Clinical Trials and Therapeutic Development**: The identification of biomarkers opens avenues for conducting clinical trials to validate their utility in clinical practice. Trials can evaluate the prognostic value of biomarkers, their role in predicting treatment response, and their ability to guide therapeutic decisions. Furthermore, biomarkers may facilitate the development and testing of new therapeutic agents aimed at modulating the dysregulated pathways identified in the study.

Research into cardiac remodeling post-ischemic injury reveals significant clinical implications, particularly regarding the disruption of cardiogenic regulators by TGF-beta and Angiotensin II. Understanding how these pathways affect critical regulators like Isl1, Nkx2–5, GATA4, and others identifies potential therapeutic targets. Drug development aimed at modulating these pathways could enhance cardiac repair and regeneration following ischemic injury. Moreover, insights into disrupted cardiogenic regulators could lead to improved diagnosis and prognosis. Biomarkers associated with these regulators may serve as diagnostic tools to assess the severity of ischemic injury and predict patient outcomes, enabling more personalized treatment approaches. This knowledge also informs surgical and interventional strategies by elucidating how remodeling alters cardiac structure and function. Optimal timing for interventions can be identified to maximize effectiveness in repairing damaged myocardium. Knowledge of pathways involved in remodeling also guides preventive measures to mitigate ischemic injury risk factors. Lifestyle interventions and medications targeting predisposing conditions could potentially reduce the impact of ischemic events on the heart. Furthermore, these insights pave the way for clinical trials evaluating new therapies targeting TGF-beta and Angiotensin II pathways. Such research may yield new treatments aimed at mitigating post-ischemic cardiac remodeling and improving patient outcomes.

## Conclusions

Cardiac remodeling following ischemic injury is a complex process influenced significantly by TGF-beta and Angiotensin II, which disrupt a multitude of cardiogenic regulators essential for cardiac development and repair. This study provides several key insights into how these signaling pathways perturb crucial regulators such as Isl1, Brg1/Baf60 – Smarcd3 complex, Nkx2–5, GATA4, Tbx5, Mef2c, HAND1/2, MYOCD, MSX2, HOPX, as well as Wnt-signaling pathway, Notch, FGF, and BMPs. We have discerned that TGF-beta and Angiotensin II exert their disruptive effects through various mechanisms, including modulation of receptor expression, interference with intracellular signaling cascades essential for cardiomyocyte differentiation and survival, induction of hypertrophic responses, and promotion of fibrosis. These disruptions collectively impair the regulatory networks that govern cardiac development and repair, leading to compromised myocardial function and structural integrity.

Understanding these mechanisms provides a comprehensive framework for developing targeted therapeutic strategies aimed at mitigating the detrimental effects of TGF-beta and Angiotensin II in ischemic heart disease. Restoration of normal cardiogenic regulator function holds promise for enhancing cardiac repair and regeneration, thereby improving clinical outcomes for patients suffering from ischemic heart injury. Ischemic injury to the heart initiates a cascade of tissue damage and triggers complex adaptive responses, leading to structural changes and functional adaptations aimed at compensating for myocardial damage. Central to these remodeling processes is the upregulation of TGF-beta signaling, which plays a crucial role in modulating cardiogenic regulators implicated in cardiac repair. TGF-beta disrupts several key regulators such as Isl1, Brg1/Baf60 – Smarcd3 complex, Nkx2–5, GATA4, Tbx5, Mef2c, HAND1/2, MYOCD, MSX2, HOPX, and pathways like Wnt, Notch, FGF, and BMPs. Mechanisms include altering receptor expression, interfering with signaling pathways such as the Smad pathway, and promoting fibrotic responses, all of which contribute to maladaptive remodeling post-ischemic injury. Angiotensin II further exacerbates cardiac remodeling following ischemic injury by disrupting similar cardiogenic regulators through hypertrophic pathways and induction of fibrosis. It alters BMP signaling and MAPK/ERK pathways, exacerbates oxidative stress, and triggers inflammatory responses, compounding the adverse effects on myocardial repair and regeneration. The interplay between TGF-beta and Angiotensin II pathways involves synergistic or antagonistic interactions that significantly impact downstream signaling pathways crucial for cardiac repair.

Understanding these interactions informs potential therapeutic strategies aimed at mitigating adverse remodeling effects and promoting cardiac recovery. Targeted therapeutic approaches could include inhibitors targeting TGF-beta and Angiotensin II signaling pathways to restore normal cardiogenic regulator function and enhance myocardial repair mechanisms post-ischemic injury. Looking ahead, future research directions involve further exploration of molecular interactions and signaling cascades involved in cardiac remodeling. Translational research efforts are essential to develop new therapies that improve outcomes in ischemic heart disease by effectively targeting the underlying mechanisms of remodeling and enhancing cardiac function and repair.

## Abbreviations

TGF-beta: Transforming Growth Factor-beta
Ang II: Angiotensin II
Isl1: Islet1
Brg1/Baf60: Brahma-related gene 1 / BAF60 - SWI/SNF-related matrix-associated actin-dependent regulator of chromatin subfamily D member 3
Nkx2-5: NK2 Homeobox 5
GATA4: GATA Binding Protein 4
Tbx5: T-Box Transcription Factor 5
Mef2c: Myocyte Enhancer Factor 2C
HAND1/2: Heart and Neural Crest Derivatives Expressed ½
MYOCD: Myocardin
MSX2: Msh Homeobox 2
HOPX: Homeodomain Only Protein X
Wnt: Wingless-Related Integration Site
Notch: Notch Receptors
FGF: Fibroblast Growth Factor
BMPs: Bone Morphogenetic Proteins
RAAS: Renin-Angiotensin-Aldosterone System
ECM: Extracellular Matrix
MAPK: Mitogen-Activated Protein Kinase

## Declarations

### Ethics declarations

#### Ethics approval and consent to participate

Not applicable.

### Consent for publication

Not applicable.

### Data Availability statement

All data generated or analyzed during this study are included in this article.

## Acknowledgements

“Not applicable”.

## Authors’ Information

**1. Ovais Shafi (OS)\*** is the author of the study and was involved in the idea, concept, design, and methodology of the study, literature search and references. He did the writing, editing, and revision of the manuscript. He was involved in drawing the findings, results, conclusions, implications of the study, interpretation of the data and was involved in all aspects of the study. He prepared and wrote discussion, results, conclusions and all areas of the study. OS extracted and analyzed the data. He was involved in critical evaluation, audit of every aspect of the study, data extraction, adherence of the study to relevant PRISMA guidelines, limitations of the study, references, and all others. He was involved in drawing fig 1. The author read and approved the manuscript.

**Fig 1.**
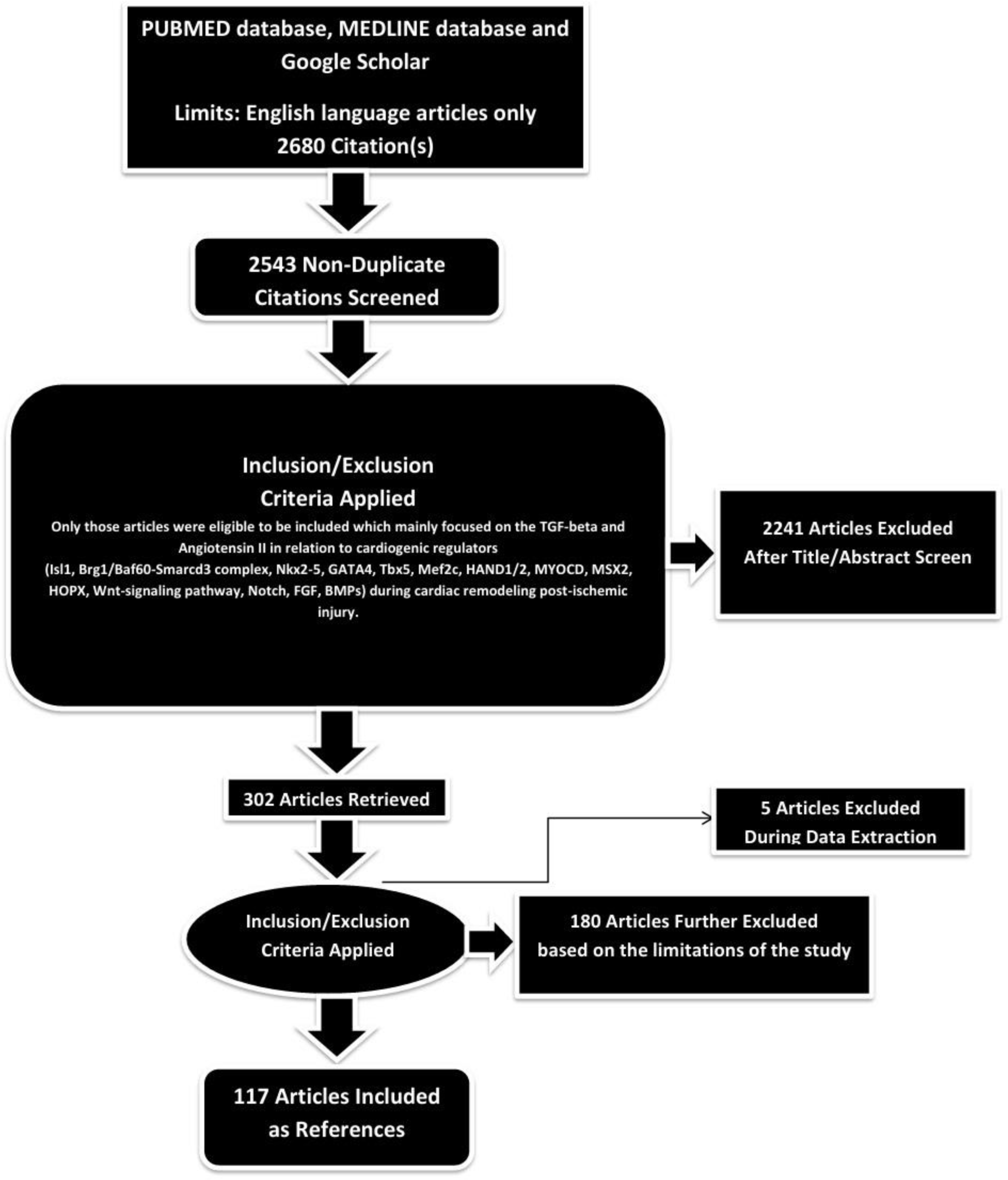
PRISMA FLOW DIAGRAM: This figure represents graphically the flow of citations in the study.

**Ovais Shafi (OS)\***, MBBS - Sindh Medical College - Dow University of Health Sciences, Karachi, Pakistan. He aspires to become an eminent ‘Physician Scientist’. He is devoted to the research in disease development mechanisms, disease origins and therapeutics. OS is also passionate about multiple research areas including clinical trials, clinical medicine, therapeutics, regenerative medicine, precision medicine including gene therapies, finding disease specific targets for gene therapy, role of disease genomics and epigenetics in diagnosis, management, and therapeutics development. He is dedicated to the field of research and clinical medicine.

**Corresponding author: OS**

Correspondence to Ovais Shafi

**2. Kashaf Zahra (KZ)** is also the author of the study and contributed to the writing, editing and revision. She also contributed to the results and conclusions sections of the study along with working on the findings, interpretation of the data and references.

Kashaf Zahra, MBBS, MD, Nishtar Medical University, Pakistan. She is passionate about research in disease origins and disease development mechanisms. One of the major challenges in the field of medicine is the lack of treatments due to unknown etiologies of diseases and due to unknown areas in pathogenesis. She is dedicated and passionate about research in this area as this area has huge potential to revolutionize the field of medicine by improving the patient outcomes. She is also passionate about translating the findings of research in disease origins/ disease development mechanisms into clinical implications. Her goals include: Healthcare provider and public service.

**3. Haider Hussain Shah (HHS)** is also the author of the study and contributed to the writing, editing and revision. He also contributed to the results and conclusions sections of the study along with working on the findings, interpretation of the data and references.

Haider Hussain Shah, MD, MBBS - Sindh Medical College - Jinnah Sindh Medical University / Dow University of Health Sciences, Karachi, Pakistan. He is currently working as Medical Hospitalist at Bayhealth Hospital - Kent Campus, Dover, Delaware, USA. He is dedicated to the field of medicine and research. His goals include: Healthcare provider and public service.

***Ovais Shafi (OS)*, Kashaf Zahra (KZ)*** *and **Haider Hussain Shah (HHS)** are the authors of this research study. The work and contributions of the authors have been described in detail. All authors have read and approved the manuscript*.

